# Amerind ancestry predicts the impact of *FADS* genetic variation on omega-3 PUFA deficiency, cardiometabolic and inflammatory risk in Hispanic populations

**DOI:** 10.1101/2021.04.16.21255626

**Authors:** Chaojie Yang, Brian Hallmark, Jin Choul Chai, Timothy D. O’Connor, Lindsay M Reynolds, Alexis C Wood, Michael Seeds, Yii-Der Ida Chen, Lyn M Steffen, Michael Y Tsai, Robert C. Kaplan, Martha L. Daviglus, Lawrence J. Mandarino, Amanda M. Fretts, Rozenn N Lemaitre, Dawn K. Coletta, Sarah A. Blomquist, Laurel M. Johnstone, Chandra Tontsch, Qibin Qi, Ingo Ruczinski, Stephen S Rich, Rasika A Mathias, Floyd H Chilton, Ani Manichaikul

## Abstract

Hispanic populations have higher rates of obesity, elevated triglycerides, and a greater prevalence of diabetes. Long chain polyunsaturated fatty acids (LC-PUFAs) and LC-PUFA metabolites have critical signaling roles that regulate dyslipidemia and inflammation. Genetic variation in the *FADS* cluster accounts for a large part of the interindividual differences in circulating and tissue levels of LC-PUFAs, with the genotypes most strongly predictive of low LC-PUFA levels at strikingly higher frequencies in Amerind (AI) ancestry populations. In this study, we examined relationships between genetic ancestry and *FADS* variation, plasma phospholipid levels of LC-PUFAs, anthropometric measures, and circulating metabolic and inflammatory biomarkers in 1,102 Hispanic American participants, representing six distinct ancestry populations from the Multi-Ethnic Study of Atherosclerosis. We demonstrate strong negative associations between AI genetic ancestry and LC-PUFA levels. The *FADS* rs174537 single nucleotide polymorphism (SNP) accounted for much of the AI ancestry effect on LC-PUFAs, especially for low levels of n-3 LC-PUFAs. Rs174537 was also strongly associated with several metabolic, inflammatory and anthropomorphic traits including circulating triglycerides (TGs) and E-selectin in MESA Hispanics. We further replicated the association with circulating TGs in two additional Hispanic cohorts: the Hispanic Community Health Study/Study of Latinos and the Arizona Insulin Resistance Registry. Our study demonstrates that Amerind ancestry provides a useful and readily available tool to identify individuals most likely to have *FADS*-related n-3 LC-PUFA deficiencies and associated cardiovascular risk.

## Introduction

Human diets in developed countries have changed dramatically over the past 75 years, leading to increased obesity, inflammation, cardio-metabolic disorders and cancer risk, possibly due to interactions between genotype with diet and other factors. Certain racial/ethnic groups carry a disproportionate burden of preventable negative outcomes and associated mortality.^1–3^ Hispanic populations represent the largest racial/ethnic US minority where, compared to non-Hispanic whites, they have higher rates of obesity^4^, poorly controlled high blood pressure^5^, and elevated circulating triglycerides (TGs)^6^, Hispanic populations also demonstrate a higher prevalence of diabetes and nonalcoholic fatty liver disease (NAFLD) than other racial/ethnic populations in the United States.^7, 8^ Hispanic Americans represent a heterogenous group with respect to ancestry, with notable differences in cultural/ lifestyle factors and disease prevalence based on country of origin. In particular, Hispanics identifying with the higher Amerind (AI)-ancestry origin have demonstrated enhanced urine albumin excretion^9^, heart failure^10^, lupus erythematosus risk^11^, and prevalence of NAFLD compared to other Hispanic populations^12^, supporting the critical need to conduct studies in these large, rapidly growing populations.

Omega-3 (n-3) and omega-6 (n-6) long chain (20-22 carbon; LC-) polyunsaturated fatty acids (PUFAs) and their metabolites play vital roles in innate immunity, energy homeostasis, brain development and cognitive function.^13–19^ LC-PUFAs are critical signaling molecules for immunity and inflammation with most evidence showing that n-3 and n-6 LC-PUFAs and their metabolic products have different and often opposing effects.^20–24^ Metabolites of the n-6 LC-PUFA arachidonic acid (ARA) typically act locally to promote inflammatory responses^25–27^, while n-3 LC-PUFAs, such as eicosapentaenoic acid (EPA) and docosahexaenoic acid (DHA) and their metabolites, have anti-inflammatory and “pro-resolution” properties^28, 29^. In addition to their effects on inflammation, circulating levels of n-3 LC-PUFAs, including EPA and DHA, are inversely associated with fasting and postprandial serum TG concentrations, largely through attenuation of hepatic very-low-density lipoprotein (VLDL)-TG production.^30, 31^ Dietary supplementation with these n-3 LC-PUFAs has been shown consistently to reduce fasting circulating TG levels and improve lipid accumulation associated with NAFLD.^32, 33^

The biosynthesis of n-3 and n-6 LC-PUFAs transpires via alternating desaturation (Δ6, Δ5, and Δ4) and elongation enzymatic steps encoded by fatty acid desaturase (*FADS)* cluster genes (*FADS1* and *FADS2*), and fatty acid elongase genes (*ELOVL2* and *ELOVL5)*, and there is a limited capacity for biosynthesis through this pathway.^34–36^ As a result, the primary dietary PUFAs that enter this pathway (linoleic acid [18:2n-6; LA], α-linolenic acid [18:3n-3; ALA], and their metabolic intermediates) compete as substrates for the desaturation and elongation steps. Additionally early studies with deuterated substrates indicated there is a saturation point where additional dietary quantities of 18 carbon dietary substrates had no effect on circulating LC-PUFA levels.^37^ These studies also estimated that conversion of dietary ALA provided 75-85% of total n-3 LC-PUFAs needed to meet daily requirements.^37^

In 1961, a major effort was initiated to reduce levels of saturated fatty acids and replace them with PUFAs in an attempt to reduce circulating LDL-cholesterol and TGs.^38–40^ This in turn led to a dramatic increase in eighteen carbon (18C-) PUFA-containing vegetable oils such as soybean, corn, and canola oils that contain high levels of n-6 LA relative n-3 ALA. It has been estimated that dietary LA increased from 2.79% to 7.21% of energy, whereas there was only a modest elevation in ingested ALA (from 0.39% to 0.72%), resulting in a ∼15:1 ratio of LA to ALA entering the LC-PUFA biosynthetic pathway and an estimated 40% reduction in total circulating n-3 LC-PUFA levels.^41^ Since LA and ALA compete for the same desaturation and elongation steps and there is a limited capacity for n-6 and n-3 LC-PUFA biosynthesis through the pathway, several human and animal studies suggested that the dramatic shift in quantities and ratios of dietary LA and ALA could lead to imbalances in n-6 to n-3 LC-PUFAs and, potentially, n-3 LC-PUFA deficiencies ^42–46^ Thus, as certain populations moved from traditional to modern Western diets (MWD), it was suggested excess LA would lead to ‘Omega-3 Deficiency Syndrome’.^47^

The rate limiting step of LC-PUFA biosynthesis has long been recognized to be the *FADS*-encoded Δ6 and Δ5 desaturation steps. Over the past decade, GWAS and candidate gene studies have shown that variation in the *FADS* gene locus on human chromosome 11 is strongly associated with plasma levels of ARA and EPA and the efficiency by which LC-PUFA precursors (18C dietary PUFAs) are metabolized to n-6 and n-3 LC-PUFAs.^48, 49^ *FADS* cluster genetic variation is associated with numerous molecular phenotypes that impact human disease as well as the risk of several diseases, including coronary heart disease^50^, diabetes^51–53^ and colorectal cancer^54^. *FADS* cluster genetic variation is strongly associated with circulating TG and VLDL concentrations in young healthy Mexicans.^55^

Our previous studies revealed that African (compared to European) ancestry populations had elevated levels of LC-PUFAs, an increased frequency of the associated *FADS* genetic variants and a more efficient LC-PUFA biosynthesis (termed the “derived” haplotype).^56^ In contrast, *FADS* variants associated with more limited capacity to synthesize LC-PUFAs (termed “ancestral” haplotype) are nearly fixed in Native American and Greenland Inuit populations and found at high frequencies in Amerind (AI) Ancestry Hispanic populations.^56^ These distinct patterns of haplotypes have resulted in part from positive selection for the ancestral haplotype among Indigenous American populations.^56^

While the role of *FADS* variation in modulating circulating fatty acid levels has been documented previously^48, 49^, prior studies have not examined the impact that population differences in *FADS* allele frequencies have in downstream population-specific risk of fatty acid deficiency, The hypothesis tested in this paper is that ancestral *FADS* variation in the context of MWD is associated with low (perhaps inadequate) circulating levels of LC-PUFAs (particularly n-3 LC-PUFAs) in a large proportion of high AI-Ancestry Hispanic populations compared to other Hispanic populations, with downstream effects on numerous cardiometabolic and inflammatory risk factors. To address this question, we first examined the relationship between the genomic proportions of AI ancestry and circulating phospholipid LC-PUFA levels in self-reported Hispanic individuals from the Multi-Ethnic Study of Atheroscloersis (MESA)^57^, which includes Hispanic groups with varying levels of AI ancestry. Second, we assessed the extent to which this relationship is explained by genetic variation within the *FADS1/2* locus, and also examined the impact of *FADS* genetic variation on cardiometabolic and inflammatory risk factors (lipids, anthropometric and inflammatory markers). Third, we tested whether these *FADS* genetic associations replicated in two high AI-Ancestry Hispanic cohorts, the Arizona Insulin Resistance (AIR) Registry^58^, and the Hispanic Community Health Study/Study of Latinos (HCHS/SOL)^59, 60^.

## Results

### Participant Characteristics

The MESA participants^57, 61^ included in this analysis comprised 1,102 unrelated individuals aged 45 to 84 years at baseline of self-reported Hispanic race/ethnicity with country-specific classification based on the birthplace of parents and grandparents corresponding to Central American (n=80), Cuban (n=45), Dominican (n=145), Mexican (n=572), Puerto Rican (n=167) and South American (n=93) **(Table 1)**. MESA Hispanic participants were recruited primarily from three field centers in the United States (Columbia University, University of California – Los Angeles (UCLA) and the University of Minnesota). The global proportions of AI, African, and European genetic ancestry in each individual were estimated using genome-wide SNP data (**Table 1**). Higher frequencies of the rs174537 T allele in the *FADS* cluster (corresponding to the ancestral allele) were observed in subjects with country/region-specific origins in Central America (0.59), South America (0.56) and Mexico (0.59) compared to those of Dominican (0.27), Cuban (0.28) or Puerto Rican origin (0.40) (**Table 1**).

**Table 1:**
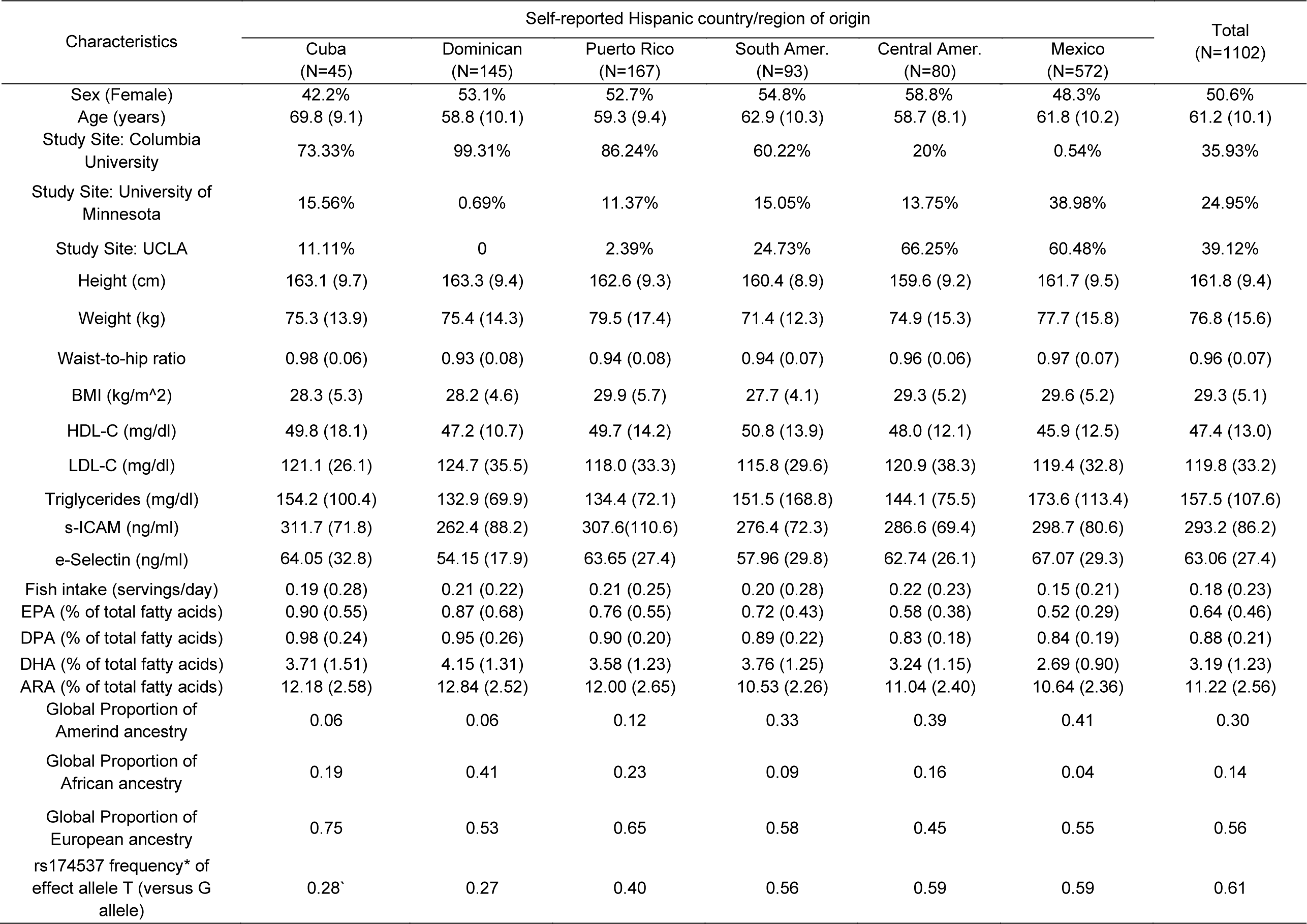
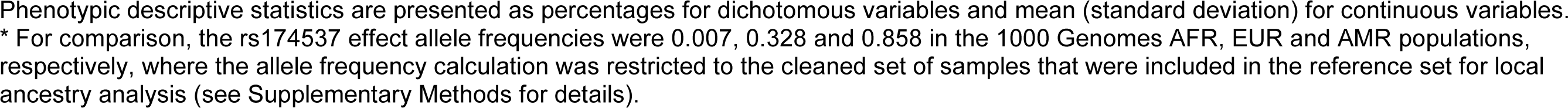
Participant Characteristics for individuals of self-identified Hispanic origin from the MESA cohort

| Characteristics | Self-reported Hispanic country/region of origin |  |  |  |  |  | Total<br>(N=1102) |
| --- | --- | --- | --- | --- | --- | --- | --- |
|  | Cuba<br>(N=45) | Dominican<br>(N=145) | Puerto Rico<br>(N=167) | South Amer.<br>(N=93) | Central Amer.<br>(N=80) | Mexico<br>(N=572) |  |
| Sex (Female) | 42.2% | 53.1% | 52.7% | 54.8% | 58.8% | 48.3% | 50.6% |
| Age (years) | 69.8 (9.1) | 58.8 (10.1) | 59.3 (9.4) | 62.9 (10.3) | 58.7 (8.1) | 61.8 (10.2) | 61.2 (10.1) |
| Study Site: Columbia University | 73.33% | 99.31% | 86.24% | 60.22% | 20% | 0.54% | 35.93% |
| Study Site: University of Minnesota | 15.56% | 0.69% | 11.37% | 15.05% | 13.75% | 38.98% | 24.95% |
| Study Site: UCLA | 11.11% | 0 | 2.39% | 24.73% | 66.25% | 60.48% | 39.12% |
| Height (cm) | 163.1 (9.7) | 163.3 (9.4) | 162.6 (9.3) | 160.4 (8.9) | 159.6 (9.2) | 161.7 (9.5) | 161.8 (9.4) |
| Weight (kg) | 75.3 (13.9) | 75.4 (14.3) | 79.5 (17.4) | 71.4 (12.3) | 74.9 (15.3) | 77.7 (15.8) | 76.8 (15.6) |
| Waist-to-hip ratio | 0.98 (0.06) | 0.93 (0.08) | 0.94 (0.08) | 0.94 (0.07) | 0.96 (0.06) | 0.97 (0.07) | 0.96 (0.07) |
| BMI (kg/m<sup>2</sup>) | 28.3 (5.3) | 28.2 (4.6) | 29.9 (5.7) | 27.7 (4.1) | 29.3 (5.2) | 29.6 (5.2) | 29.3 (5.1) |
| HDL-C (mg/dl) | 49.8 (18.1) | 47.2 (10.7) | 49.7 (14.2) | 50.8 (13.9) | 48.0 (12.1) | 45.9 (12.5) | 47.4 (13.0) |
| LDL-C (mg/dl) | 121.1 (26.1) | 124.7 (35.5) | 118.0 (33.3) | 115.8 (29.6) | 120.9 (38.3) | 119.4 (32.8) | 119.8 (33.2) |
| Triglycerides (mg/dl) | 154.2 (100.4) | 132.9 (69.9) | 134.4 (72.1) | 151.5 (168.8) | 144.1 (75.5) | 173.6 (113.4) | 157.5 (107.6) |
| s-ICAM (ng/ml) | 311.7 (71.8) | 262.4 (88.2) | 307.6(110.6) | 276.4 (72.3) | 286.6 (69.4) | 298.7 (80.6) | 293.2 (86.2) |
| e-Selectin (ng/ml) | 64.05 (32.8) | 54.15 (17.9) | 63.65 (27.4) | 57.96 (29.8) | 62.74 (26.1) | 67.07 (29.3) | 63.06 (27.4) |
| Fish intake (servings/day) | 0.19 (0.28) | 0.21 (0.22) | 0.21 (0.25) | 0.20 (0.28) | 0.22 (0.23) | 0.15 (0.21) | 0.18 (0.23) |
| EPA (% of total fatty acids) | 0.90 (0.55) | 0.87 (0.68) | 0.76 (0.55) | 0.72 (0.43) | 0.58 (0.38) | 0.52 (0.29) | 0.64 (0.46) |
| DPA (% of total fatty acids) | 0.98 (0.24) | 0.95 (0.26) | 0.90 (0.20) | 0.89 (0.22) | 0.83 (0.18) | 0.84 (0.19) | 0.88 (0.21) |
| DHA (% of total fatty acids) | 3.71 (1.51) | 4.15 (1.31) | 3.58 (1.23) | 3.76 (1.25) | 3.24 (1.15) | 2.69 (0.90) | 3.19 (1.23) |
| ARA (% of total fatty acids) | 12.18 (2.58) | 12.84 (2.52) | 12.00 (2.65) | 10.53 (2.26) | 11.04 (2.40) | 10.64 (2.36) | 11.22 (2.56) |
| Global Proportion of Amerind ancestry | 0.06 | 0.06 | 0.12 | 0.33 | 0.39 | 0.41 | 0.30 |
| Global Proportion of African ancestry | 0.19 | 0.41 | 0.23 | 0.09 | 0.16 | 0.04 | 0.14 |
| Global Proportion of European ancestry | 0.75 | 0.53 | 0.65 | 0.58 | 0.45 | 0.55 | 0.56 |
| rs174537 frequency* of effect allele T (versus G allele) | 0.28 | 0.27 | 0.40 | 0.56 | 0.59 | 0.59 | 0.61 |

Phenotypic descriptive statistics are presented as percentages for dichotomous variables and mean (standard deviation) for continuous variables. \* For comparison, the rs174537 effect allele frequencies were 0.007, 0.328 and 0.858 in the 1000 Genomes AFR, EUR and AMR populations, respectively, where the allele frequency calculation was restricted to the cleaned set of samples that were included in the reference set for local ancestry analysis (see Supplementary Methods for details).

### LC-PUFA levels are associated with Amerind genetic ancestry

Higher proportions of AI genomic ancestry were associated with lower levels of LC-PUFAs in MESA Hispanics participants. **Figure 1 (left panel)** shows levels of EPA, DHA and ARA (expressed as the percentage of total fatty acids here and throughout the entire manuscript) as a function of inferred AI ancestry. Overall, AI ancestry explained 12.32%, 12.30% and 12.48% of total variation in EPA, DHA and ARA, respectively. Each 10% increase in AI ancestry was associated with a decrease of EPA (0.049), DHA (0.185) and ARA (0.401) in phospholipids. Between subjects with the lowest and highest proportions of AI ancestry, the n-3 LC-PUFAs decreased by 60.6% (for EPA) and 46.8% (for DHA) and the n-6 LC-PUFAs decreased by 30.7% (for ARA). Consequently, the nadir in predicted fatty acids levels in plasma phospholipids among those with 100% AI ancestry was ∼0.3 and ∼2 for EPA and DHA, respectively, compared to ∼8.6 for the n-6 LC-PUFA, ARA.

**Figure 1.**
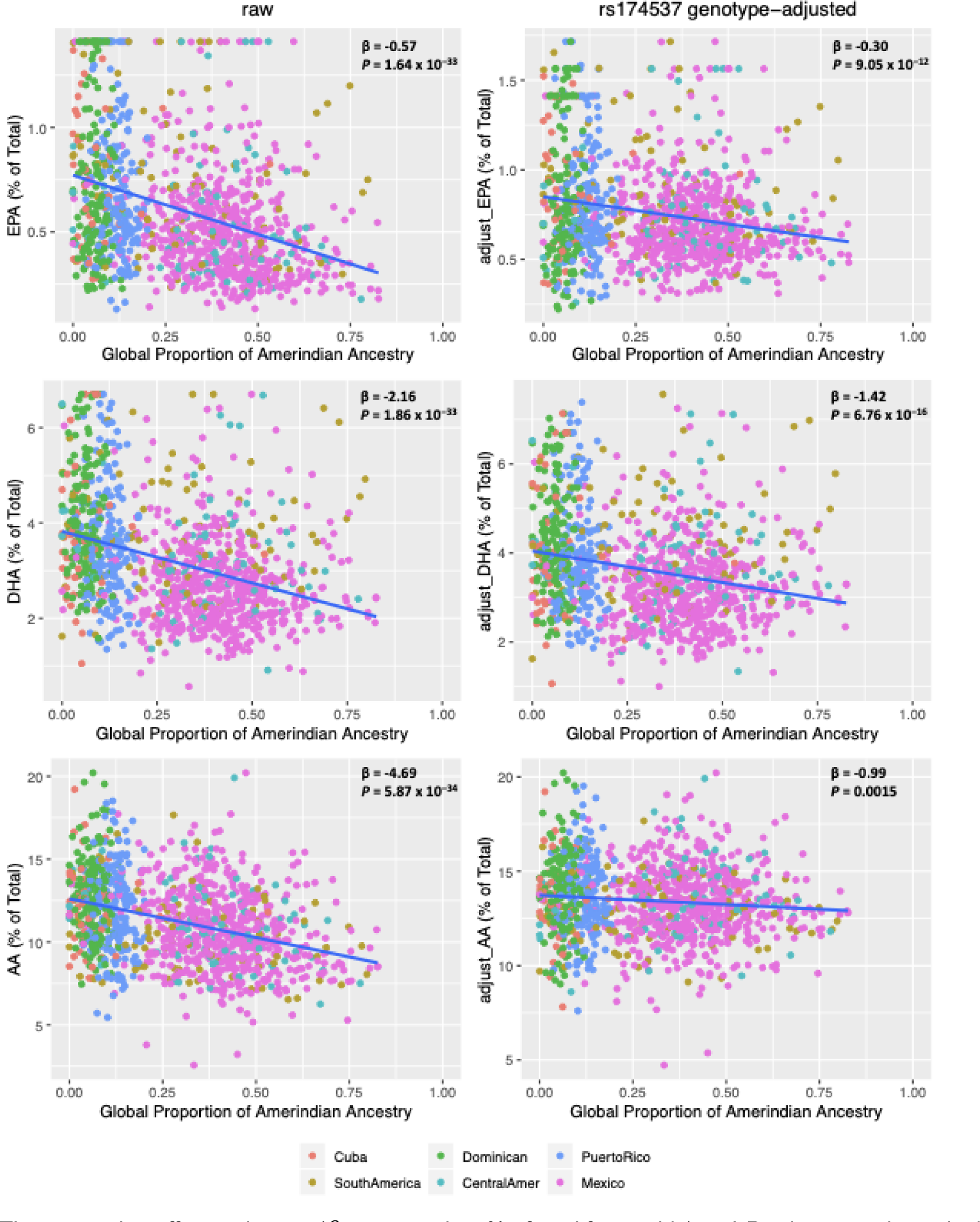
Relationship of LC-PUFA levels with Global Proportion of Amerind Ancestry before and after adjustment for rs174537 genotype. The regression effect estimates (β expressed as % of total fatty acids) and *P*-values are shown in the upper right corner of each panel. **Left** panels show the relationship of raw LC-PUFA levels with Global Proportion of Amerind Ancestry as estimated from genome-wide SNP data. **Right** panels show the relationship of rs174537 genotype-adjusted LC-PUFA levels with Global Proportion of Amerind Ancestry. Here, the adjusted LC-PUFA levels were obtained as residuals after regression against rs174537 genotype and re-centered around the raw mean.

Given the prior evidence that key genetic determinants of LC-PUFAs mapping to the *FADS* locus show strong variation in frequency between populations, we sought to determine the role of *FADS* variation in the relationships between LC-PUFA levels and global AI ancestry. LC-PUFAs were adjusted for rs174537 genotype (**Figure 1, right panels);** rs174537 is selected as a representative proxy SNP for the well documented associations between the *FADS* locus and LC-PUFAs^62, 63^. The rs174537 SNP has a strong effect on the ancestry-related decline in all LC-PUFAs. After adjusting for rs174537 genotype, an inverse association remains between global proportion of AI ancestry and EPA (β = -0.30, 95% Confidence Interval [Ci] = [-0.39, -0.22], *P* = 9.05 x 10^-12^), DHA (β = -1.42, 95% CI = [-1.76, -1.08], *P* = 6.76 x 10^-16^) and ARA (β = -0.99, 95% CI = [-1.59, -0.38], *P* = 0.0015). Regression analysis of n-3 and n-6 LC-PUFAs with global proportion of AI ancestry, accounting for covariates: age, sex and fish intake (Model 1), resulted in inverse relationships between the global proportion of AI ancestry with EPA (β = -0.48, *P* = 3.7 x 10^-23^), DPA (β = -0.18, *P* = 7.6 x 10^-06^), DHA (β = -0.63, *P* = 0.0007) and ARA (β = -4.06, *P* = 1.3 x 10^-16^) **(Table S1).** These effects were consistent across study sites in MESA, with the largest effects observed at the University of Minnesota field center (**Table S1**). Accounting for rs174537 genotype (Model 2), there remained an inverse association between the global proportion of AI ancestry with EPA (β = -0.28, *P* = 3.7 x 10^-08^) (**Table S1**), while the relationship of global proportion of AI ancestry with ARA, DPA and DHA was no longer significant (**Table S1**). In a model further accounting for local AI ancestry in addition to rs174537 (Model 3), EPA continued to be inversely associated with global proportion of AI ancestry (β = -0.34, *P* = 8.4 x 10^-07^), while the associations with DPA, DHA and ARA were not statistically significant (**Table S1**). In additional analysis examining global and local ancestry as potential modifiers of the effect of rs174537 on circulating fatty acid levels, we did not observe statistically significant evidence of interaction **(Table S2).**

**Table 2.** Genotypic effects of rs174537 on Fasting Lipids, Anthropometrics and inflammatory.

|  |  |  | Beta | P value |
| --- | --- | --- | --- | --- |
| Fasting Lipids | Triglycerides (mg/dL) | GT | 21.27 | 0.0001 |
|  |  | TT | 29.94 | 1.01 x 10<sup>-06</sup> |
|  | HDL-C (mg/dL) | GT | -1.30 | 0.141 |
|  |  | TT | -2.48 | 0.010 |
| Anthropometrics | waist-hip ratio | GT | 0.006 | 0.152 |
|  |  | TT | 0.013 | 8.94 x 10<sup>-03</sup> |
|  | Height (cm) | GT | -1.36 | 0.002 |
|  |  | TT | -3.46 | 6.59 x 10<sup>-12</sup> |
|  | Weight (kg) | GT | -1.89 | 0.077 |
|  |  | TT | -3.12 | 7.48 x 10<sup>-03</sup> |
|  | BMI (kg/m<sup>2</sup>) | GT | -0.25 | 0.50 |
|  |  | TT | -0.002 | 0.99 |
| Inflammatory | s-ICAM (ng/mL) | GT | 30.64 | 0.002 |
|  |  | TT | 26.09 | 0.018 |
|  | e-Selectin (ng/mL) | GT | 10.00 | 0.048 |
|  |  | TT | 11.50 | 0.032 |

**Table 2** shows the regression analysis results for the effect of rs174537 genotype on Fasting Lipids, Anthropometrics and inflammatory with adjustment for age and sex. For the effect sizes, the effects of GT and TT are in reference to GG.

As other studies have suggested different specific variants as potentially functional within the *FADS* region, we further repeated the analysis presented in **Figure 1** through sensitivity analysis focused on the *FADS* region variant rs174557 **(Figure S1)**, a common variant that diminishes binding of *PATZ1*, a transcription factor conferring allele-specific downregulation of FADS1.^64^ After adjusting for rs174557 genotype, we observed association between global proportion of AI ancestry and LC-PUFA levels (EPA: β = -0.30, *P* = 2.19 x 10^-11^; DHA: β = -1.39, *P* = 2.54 x 10^-15^; ARA: β = -1.02, *P* = 0.0012) similar to that seen after adjusting for rs174537.

### Association of global Amerind ancestry with triglycerides

Higher global proportions of AI ancestry were significantly associated with higher levels of circulating triglycerides (TG) in MESA Hispanic participants (β = 65.40 mg/dL, 95% CI = [42.28, 88.52] *P =* 3.58 x 10^-8^) **(Figure 2 -left).** This relationship was attenuated after adjusting for rs174537 (**Figure 2 -right**), although there remained a significant relationship between global proportions of AI ancestry and TG levels (β = 39.47 mg/dL, 95% CI = ([2.62, 52.05], *P =* 8.16 x 10^-04^). In sensitivity analysis, circulating triglycerides (TG) were adjusted for the variant rs174557.

**Figure 2.**
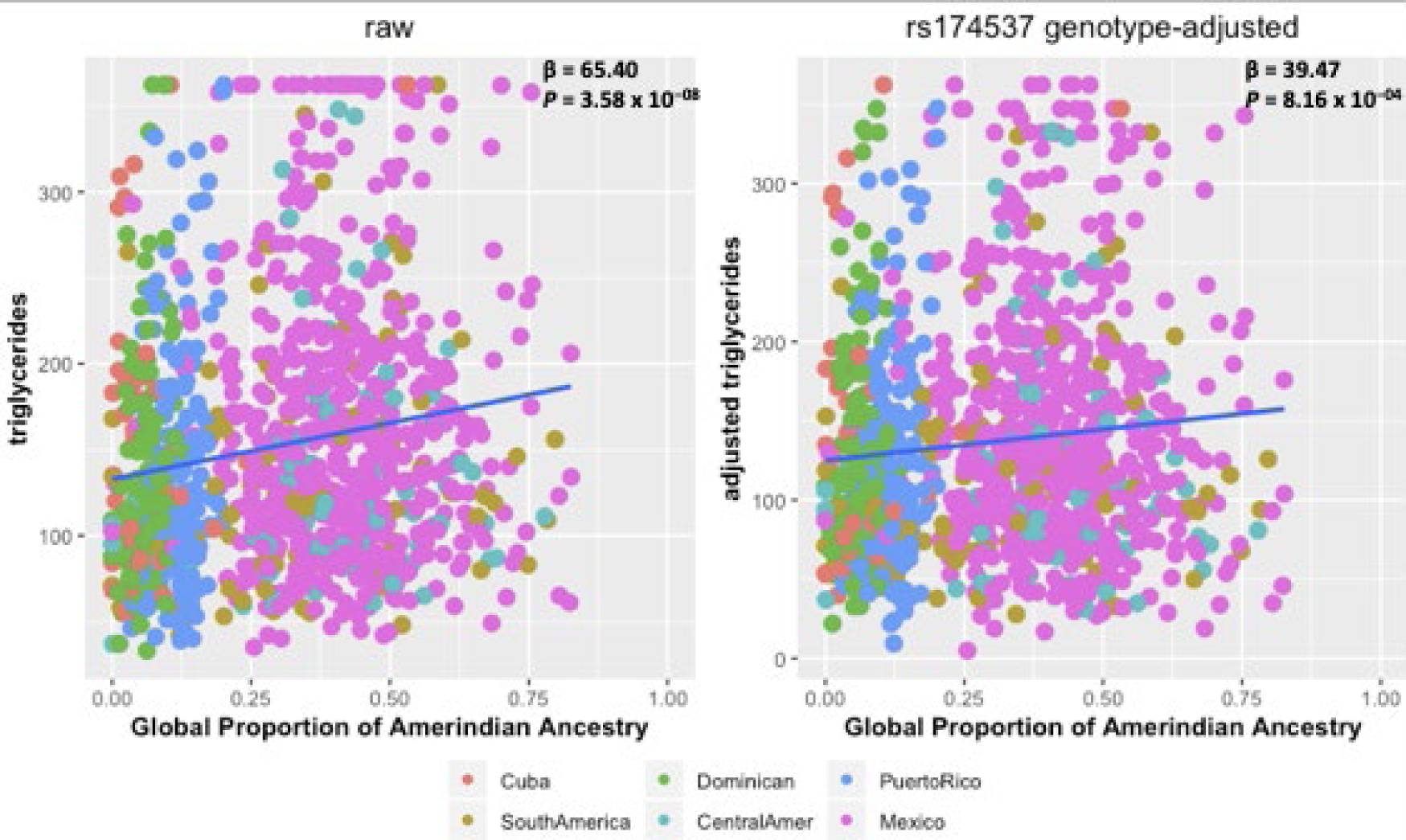
Relationship of triglycerides with Global Proportion of Amerind Ancestry before and after adjustment for rs174537 genotype. The regression effect estimates (β in mg/dL) and *P*-values are shown in the upper right corner of each panel. **Left** figure shows the relationship of raw triglyceride levels with Global Proportion of Amerind Ancestry. **Right** figure shows the relationship of rs174537 genotype-adjusted triglyceride levels with Global Proportion of Amerind Ancestry. Here, rs174537 genotype-adjusted triglyceride levels were obtained as residuals from regression accounting for rs174537 genotype, and re-centered around the raw means.

The relationship between global proportion of AI ancestry and TG levels (**Figure S2**; β = 38.93 mg/dL, *P =* 9.59 x 10^-04^) is similar with the association adjusting for rs174537. After adjusting for age and sex, the rs174537 T allele was significantly associated with higher levels of TG (GT vs GG: β = 21.27 mg/dL, 95% CI = [10.29, 32.25], *P =* 0.0001, TT vs GG: β = 29.94 mg/dL, 95% CI = [17.98, 41.88], *P =* 1.01 x 10^-6^) **(Table 2 and Figure 3)**.

**Figure 3:**
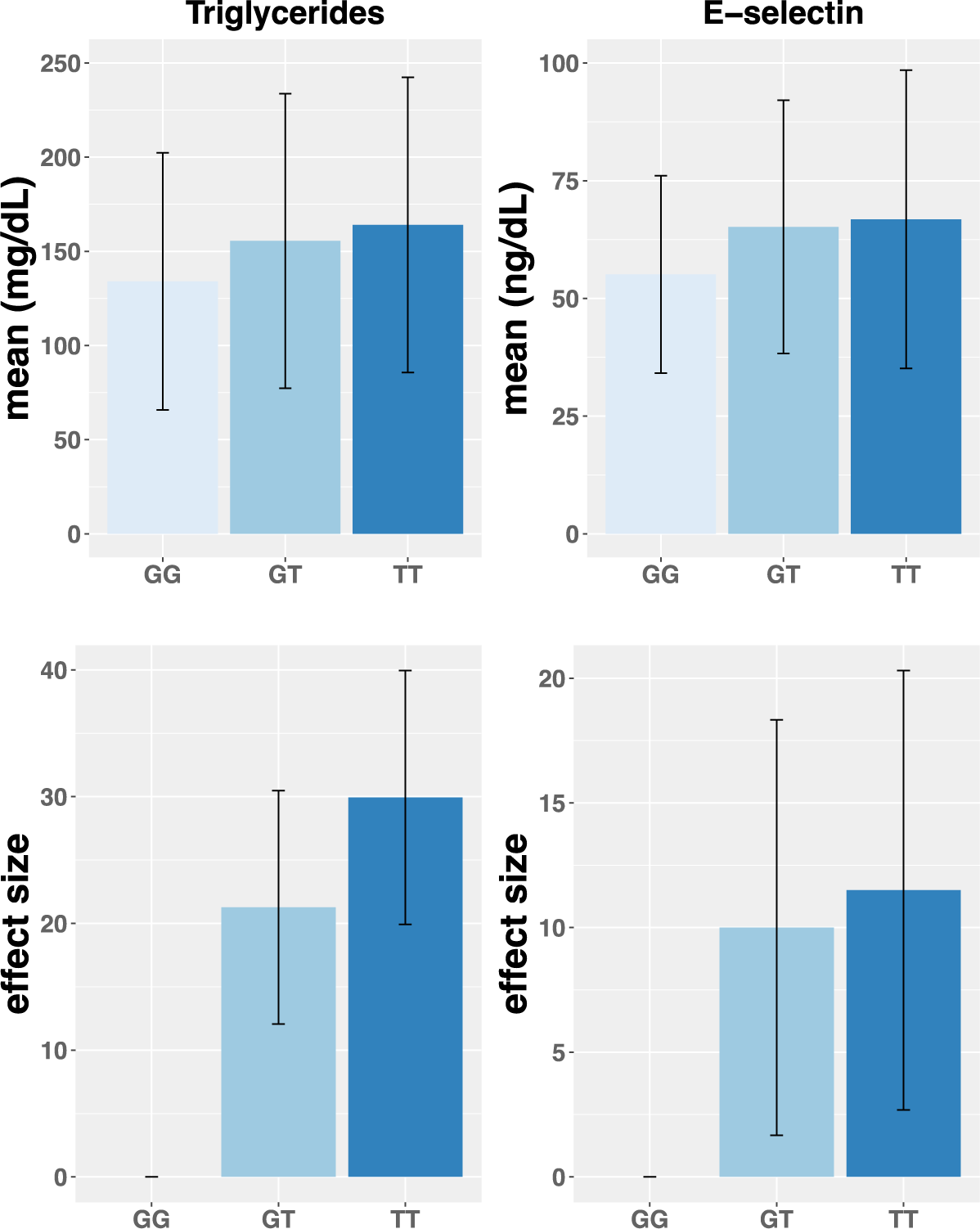
Genotypic effects of rs174537 on Triglycerides and E-selectin. Figure 3 shows the effect of rs174537 on Triglycerides and E-selectin. The sample size across genotype is 293 for GG, 484 for GT, 325 for TT. Upper figure shows the mean and standard deviation of Triglycerides and E-selectin stratified by rs174537 genotypes and lower figure shows the estimated effect and standard error of carrying one or two copies of the ancestral allele (compared to the reference of zero), after covariate-adjustment for age and sex.

### Association of PUFAs with *FADS cluster* SNPs

We performed an association analysis adjusting for rs174537 genotype to determine if there was any residual association in the *FADS* region in the MESA participants. In each of the Hispanic subgroups, after accounting for the rs174537 SNP, no additional genetic variants in the region were associated with EPA, DPA, DHA or ARA **(Figure S3**). The rs174537 SNP is in strong linkage disequilibrium with other *FADS* cluster SNPs; thus, subsequent analyses are focused solely on the rs174537 SNP.

### Effects of rs174537 on Inflammatory Biomarkers, Fasting Lipids and Anthropometrics

The effect of the *FADS* cluster SNP rs174537 on height, weight, body mass index (BMI), waist-hip ratio, s-ICAM, e-Selectin and HDL-C was estimated in the MESA Hispanic participants. In a model adjusted for age and sex, the rs174537 T allele was significantly associated with lower levels of HDL-C, higher waist-hip, lower height and weight, and higher levels of the inflammatory markers e-Selectin and s-ICAM (**Table 2, Figure 3 and Table S3**). In regression analysis with adjustment for principal components of ancestry, the rs174537 T allele remained significantly associated with higher TGs and lower height, while the associations with weight, waist-hip ratio, s-ICAM, e-Selectin and HDL-C were no longer statistically significant (**Figure S4 and Table S3**). In sensitivity analysis, we also examined the effect of rs174557 on the same set of phenotypes as examined for rs174537. Similar to the rs174537 T allele, the rs174557 A allele was significantly associated with lower levels of HDL-C, higher waist-hip, lower height and weight, and higher levels of the inflammatory markers s-ICAM **(Table S4 and Figure S4).** In regression analysis with adjustment for principal components of ancestry, the rs174557 A allele remained significantly associated with higher TGs and lower height, while the associations with weight, waist-hip ratio, s-ICAM, e-Selectin and HDL-C were no longer statistically significant **(Table S4)**.

### Replication in the AIR registry and HCHS/SOL cohort

We conducted analyses in the AIR registry (n = 497) and HCHS/SOL (n = 12,333) cohorts to examine the genotypic effect of rs174537 on multiple phenotypic traits including TGs and waist-to-hip ratio (**Table S5 and S6**). In regression analyses adjusted for age and sex (and inclusion of random effects for household block and unit sharing in HCHS/SOL), the rs174537 T allele was significantly associated with TGs (AIR: β = 10.4 mg/dL, *P* = 0.03, HCHS/SOL: β = 8.75 mg/dL, *P =* 5.84 x 10^-25^) (**Table S7**). The rs174537 T allele was also significantly associated with reduced height (β = -1.33, *P =* 4.47 x 10^-56^) and weight (β = -1.25, *P =* 2.61 x 10^-08^), and increased waist-to-hip ratio (β = 0.003; *P =* 2.77 x 10^-05^) in the HCHS/SOL cohort. The direction of effect was consistent, but not statistically significant, in the much smaller AIR cohort (**Table S7**). The association of rs174537 with TGs remained statistically significant after adjustment for principal components of ancestry (β = 4.05 mg/dL, *P =* 1.26 x 10^-05^) and the effects were consistent across the HCHS/SOL study sites (**Table S8**). We did not replicate these findings in the smaller AIR registry (**Table S7**). S-ICAM and e-Selectin were not measured in either AIR or HCHS/SOL and thus could not be evaluated for replication of the findings from MESA.

## Discussion

While prior studies have identified genetic variants within the FADS locus with strong impact on fatty acid levels^48, 49^, prior literature has not examined directly the impact of population differences in allele frequencies on population-specific risk of fatty acid deficiency. In light of dramatic differences in genetic variation within the *FADS* locus across worldwide populations^56^ and the marked changes in dietary n-6 and n-3 PUFA levels and ratios over the past 75 years, we carried out a study of to examine genomic proportion of AI ancestry as a predictor of n-3 and n-6 LC-PUFA levels and related cardiometabolic and inflammatory risk in the Hispanic participants from MESA. Our study first illustrates that certain Hispanic populations and particularly high AI-Ancestry populations have high frequencies of the ancestral allele at T at rs174537. Importantly, the frequency of the TT genotype associated with limited LC-PUFA biosynthesis ranges from <1% in African-Ancestry populations including African Americans to 40-55% in high AI-Ancestry Hispanics, and ∼11% in European-Ancestry populations.^65^ In light of high ancestral frequencies in certain Hispanic populations together with elevated dietary n-6 (LA) to n-3 (ALA) PUFAs ratios (>10:1) from the MWD entering the pathway, we postulated that these populations would be most likely to saturate their capacity to synthesize LC-PUFAs and particularly n-3 LC-PUFAs. Our statistical analyses demonstrated that global proportion of AI ancestry is predictive of reduced LC-PUFA phospholipid levels in the Hispanic population of the United States, accounting for ∼12% of total variation in EPA, DHA and ARA. Further, we showed that this relationship can be explained in large part by genetic variation within the *FADS* cluster. Given that many Hispanic individuals will have reasonable knowledge of their AI ancestry, our work suggests a practical way to identify individuals likely to carry the homozygous TT genotypes, and for whom follow-up *FADS* genotyping assays may be warranted.

While both n-6 and n-3 LC-PUFAs are impacted, relatively high levels of ARA (∼8.6% of total fatty acids) remain in circulating phospholipids in even the highest AI-Ancestry populations. In contrast, n-3 LC-PUFAs including EPA and DHA are reduced to the low (perhaps inadequate) levels of ∼0.3% [EPA] and ∼2% [DHA] of total fatty acids in circulating phospholipids in high AI-Ancestry individuals. It is not possible to say with certainty what levels of EPA and DHA or ratio of EPA + DHA/ ARA would be inadequate (deficient) and have pathophysiologic impact, but these are certainly quantitatively very low concentrations and ratios of n-3 LC-PUFAs. It has been recognized that high levels of dietary LA relative to ALA from the modern Western diets (MWD) entering the LC-PUFA biosynthetic pathway are reciprocally related to levels of n-3 LC-PUFAs due to substate saturation of the enzymatic pathway.^66, 67^ Such a scenario was proposed by both Okuyama and colleagues and Lands and colleagues three decades ago to give rise to “Omega-3 Deficiency Syndrome” and “chronic pathophysiological events”.^20, 47, 68^ We propose that a limited LC-PUFA synthetic capacity in a greater proportion of AI-Ancestry Hispanics (due to the ancestral haplotype) in the context of excess dietary LA levels and high LA/ALA ratios renders inadequate n-3 LC-PUFAs more likely in this population.

Our study also suggests that *FADS* variation has large effects on some critical cardiometabolic and inflammatory risk factors. Specifically, the proportion of AI ancestry was positively related to levels of circulating TGs and much of this effect was explained by variation in the *FADS* locus. While other studies have found associations between numerous genetic loci including *FADS* SNPs and circulating TGs^69–78^, the high frequency of the ancestral *FADS* alleles (associated with elevated TGs) and their effect size in AI-Ancestry Hispanic populations that suggest that *FADS* variation is particularly relevant to TG levels in this population. The presence of the T allele at rs174537 had a large effect on circulating TG (GT vs GG: β = 21.27 mg/dL, *P =* 0.0002, TT vs GG: β = 29.94 mg/dL, *P =* 1.01 x 10^-6^) and this genotypic effect was replicated in both the AIR registry and HCHS/SOL cohort. Circulating TG are primarily synthesized in the liver and deficiencies of n-3 LC-PUFAs and imbalances of n-6 relative to n-3 PUFAs have been associated with elevated TGs and NAFLD.^79^ Elevating n-3 LC-PUFA by diet or supplementation reduces TG by promoting hepatic fatty acid oxidation and reducing synthesis (via reducing *de novo* lipogenesis and decreasing fatty acid and adipokine release from adipocytes).^80–82^ These current data suggest that inadequate levels of n-3 LC-PUFAs in AI-Ancestry Hispanic populations may impact TG formation in the liver resulting in higher levels of circulating TG and potentially NAFLD.

Waist-to-hip ratio, used to describe the distribution of body fat, has been shown to be closely associated with hypertension, diabetes, dyslipidemia and cardiovascular disease.^83^ A previous study examined genetic loci associated with BMI and waist-to-hip ratio and found nine BMI and seven central adiposity loci in Hispanic women.^84^ To date, variation within *FADS* has not been associated with waist-to-hip ratio. While our study demonstrated that the ancestral rs174537 T allele was strongly associated with a higher waist-to-hip ratio and this risk factor was replicated in HCHS/SOL, the relationship was not statistically significant after adjusting for principal components of ancestry. Thus, waist-hip-ratio is an example of a trait for which the association with AI-Ancestry is not explained in large part by *FADS* variation.

The rs174537 allele T further demonstrated association with reduced height and weight in the large HCHS/SOL cohort (n = 12,333). Fumagalli and colleagues examined indigenous Greenland Inuit and found strong signals of natural selection within the *FADS* cluster.^85^ The identified *FADS* variants were also strongly associated with anthropometric traits including body weight and height in the Inuit, and those associations were replicated in Europeans.

A wide variety of biomarkers of inflammation were measured in MESA, and there was a strong association between rs174537 and E-selectin which maintained suggestive evidence of association even after adjustment for population structure using genetic principal components of ancestry. E-Selectin (CD-62E) plays a pivotal role in the activation and adhesion of the migrating leukocytes to the endothelium.^86^ These membrane bound adhesion molecules also undergo proteolytic cleavage that generate soluble forms that can be measured in the blood.^87^ Serum levels of E-Selectin increase in many pathologies involving chronic inflammation including obesity^88^, cardiovascular disease^89^, bronchial asthma^90^ and cancer^91, 92^.

Limitations of the study include a focus on primarily urban Hispanic American populations represented by the MESA cohort, potential confounding by diet and lifestyle habits across the six Hispanic subgroups in MESA, and systematic differences in PUFA levels across MESA study sites. To address the observable variation across Hispanic subgroups and study site, we included additional analyses stratified by these factors and demonstrated that our results were consistent across strata. Additionally, we used food frequency questionnaire data to confirm participants included in our analyses did not have self-reported use of fish oil supplements, and we performed analyses adjusted for self-reported fish intake in MESA. Still, we recognize there are inherent limitations with the quality of self-report-based measures of diet and supplement use. Further, we did not consider additional measures of dietary intake of n-3 and n-6 PUFAs in our regression analyses, in part because we determined that we did not have reliable measures available for these parameters in the MESA participants. Therefore, future studies should examine further the impact of dietary differences on the relationship between AI ancestry, *FADS* variation, and LC-PUFA levels.

Despite these limitations, our study reveals that *FADS* variation in AI-Ancestry Hispanic populations is inversely associated with dyslipidemia and inflammation, risk factors for a wide range of pathologies including cardiovascular and metabolic diseases. These associations are observed strongly in these Hispanic populations in part because of the high frequencies of ancestral *FADS* alleles. It may be that LC-PUFAs or their metabolites (eicosanoids, docosanoids, resolvins, protectins, etc.) are responsible for these genetic effects given the direct relationship between *FADS* variation and LC-PUFA levels. Alternatively, we have recently combined genetic and metabolomic analyses to identify the *FADS* locus as a central control point for biologically-active LC-PUFA-containing complex lipids that act as signaling molecules such as the endocannabinoid, 2-AG, and such endocannabinoids are known to impact anthropomorphic and other phenotypic characteristics.^93^

Our results also suggest that targeting recommendations for n-3 and n-6 LC-PUFA intake/supplementation within AI-Ancestry Hispanic populations may be particularly effective. This premise is supported by the fact that numerous mechanistic studies directly link low levels of n-3 LC-PUFAs and high n-6 to n-3 ratios to elevated tissue and circulating TGs and NAFLD, and several recent reviews and meta-analyses suggest that n-3 LC-PUFA supplementation improves circulating and tissue levels of TG and NAFLD.^94, 95^ Prior research demonstrates that mean proportions of Amerind ancestry vary greatly by self-identified regions of origin among Hispanic Americans, with Mexican, Central American and South American Hispanics showing the greatest proportions, and individuals identifying as Cuban, Dominican and Puerto Rican showing considerably lower proportions.^61, 96^ While a long term goal of applying precision nutrition may include genotyping of rs174537 (or related *FADS* region variants)_in routine health care screening, current health care practice does not provide adequate resources to genotype most individuals. Thus, a priori information predictive of ancestry such as country or origin or otherwise, may serve as a preliminary tool to prioritize those who are most likely to have low circulating and tissue levels of n-3 LC-PUFA and would benefit from additional screening either through genotyping or screening for n-3 LC-PUFA deficiency. Despite the current limitations of precision nutrition including adequate genetic testing, the translational implications of this work are to point out that a large proportion of AI-Ancestry Hispanic populations have low (perhaps deficient) levels of n-3 LC-PUFAs and increased related risk factors. Thus, because of *FADS*-related deficiencies, these populations may be particularly responsive to diets or supplements enriched in n-3 LC-PUFAs.

## Methods

### Study participants

MESA is a longitudinal cohort study of subclinical cardiovascular disease and risk factors that predict progression to clinically overt cardiovascular disease or progression of subclinical disease ^57^. Between 2000 and 2002, MESA recruited 6,814 men and women 45 to 84 years of age from Forsyth County, North Carolina; New York City; Baltimore; St. Paul, Minnesota; Chicago; and Los Angeles. Participants at baseline were 38% White, 28% African American, 22% Hispanic and 12% Asian (primarily Chinese) ancestry. This manuscript focuses on Hispanic American participants from MESA. Among the MESA Hispanic participants, self-reported birthplaces for parents’ and grandparents’ country/region of origin were used to assign country/region of origin to the following categories Central America, Cuba, the Dominican Republic, Mexico, Puerto Rico and South American origin were assigned for the MESA Hispanic participants.

### Ethical review

MESA participants were consented for participation at their respective MESA study sites, and the MESA study was also reviewed and approved by the Institutional Review Boards (IRBs) at each of the participating study sites. The current investigation including activities for analysis of LC-PUFA levels in MESA was reviewed and approved by the Institutional Review Board (IRB) at the University of Virginia.

### Fatty Acid measurements

The fatty acids were measured in EDTA plasma, frozen at –70°C, using methods previously described by Cao.^97^ Here, we focus on the following n-3 and n-6 fatty acids: eicosapentaenoic acid (EPA), docosapentaenoic acid (DPA), docosahexaenoic acid (DHA), and arachidonic acid (ARA). Details of measurement and treatment of outliers are provided in the **Supplementary Methods**.

### Additional phenotypes in MESA

We considered additional phenotypes in analysis of the MESA data including lipids (HDL-C and triglycerides), anthropometric (height, weight, waist-hip ratio), and inflammatory markers (soluble e-Selectin and soluble ICAM-1). Details of measurement and treatment of outliers are provided in the **Supplementary Methods**.

### Genotyping, genetic association and ancestry analysis

Participants in the MESA cohort who consented to genetic analyses and data sharing (dbGaP) were genotyped using the Affymetrix Human SNP Array 6.0 (GWAS array) as part of the NHLBI CARe (Candidate gene Association Resource) and SHARe (SNP Health Association Resource) projects. After genotype quality control, we performed genome-wide imputation to the 1,000 Genomes Phase 3 integrated variant set.

Prior studies have highlighted multiple different *FADS* variants for their role in regulation of fatty acid synthesis, including rs174537^48^ and rs174557^64^; however, the relevant variants at the primary signal within the *FADS* region exhibit extended linkage disequilibrium across the region.^98^ Therefore, we focused our genetic analyses primarily on the variant rs174537, with additional sensitivity analyses using similar models for the variant rs174557. Imputed genotype data were used for genetic association analysis of the rs174537 and rs174557 SNPs (for which the imputation R-squared in MESA Hispanics were both 0.99). Principal components of ancestry were computed using genome-wide genotype data, as described previously.^99^ Global proportions of Amerind ancestry were estimated in MESA participants by leveraging reference samples from the 1000 Genomes^100^ and the Human Genome Diversity Project (HGDP)^101, 102^. Local ancestry for each individual was defined as the genetic ancestry at the position of *FADS* SNP rs174537, where each individual can have 0, 1 or 2 copies of an allele derived from each of the three possible ancestral populations (European, African and Amerind). Local ancestry, was estimated using the RFMix package.^103^ Details are provided in the **Supplementary Methods**.

### Regression modeling of n-3 and n-6 PUFAs

As we observed a strong effect of study site in regression analysis of all LC-PUFAs (**Table S9**), we performed regression analyses stratified by study site and combined by inverse-variance weighted meta-analysis. In order to examine the effect of global Amerind ancestry on the levels of n-3 and n-6 PUFAs in the MESA Hispanic participants, we carried out linear regression analyses using three different models for each of the PUFA levels as follows:

1. PUFA ∼ age + sex + fish intake + global proportion of Amerind ancestry
2. PUFA ∼ age + sex + fish intake + global proportion of Amerind ancestry + rs174537 genotype
3. PUFA ∼ age + sex + fish intake + global proportion of Amerind ancestry + rs174537 genotype + *FADS* region local proportion of Amerind ancestry

### Regression modeling for genotypic effects of *FADS* cluster SNP rs174537 on proximal traits

To examine the effect of *FADS* SNP rs174537 on lipids (HDL-C and triglycerides), anthropometric (height, weight, and waist-to-hip ratio), and inflammatory markers (s-ICAM and e-Selectin) in MESA Hispanic participants, we performed linear regression analysis with covariate adjustment for (1) age and sex, and (2) age, sex and the first four principal components of ancestry.

### Replication analysis in the AIR registry and HCHS/SOL cohort

We conducted follow-up regression analyses to examine the association of rs174537 with phenotypic traits in both the AIR registry and the HCHS/SOL cohort. The variant rs174537 was genotyped directly in both AIR and HCHS//SOL. Details are provided in the **Supplementary Methods**.

### Data availability

Genome-wide genotype data for the Multi-Ethnic Study of Atheroslerosis (MESA) and the Hispanic Community Health Study / Study of Latinos (HCHS/SOL) are available through dbGaP. The dbGaP accession numbers are: MESA phs000209 and HCHS/SOL phs000810.

## Data Availability

Genome-wide genotype data for the Multi-Ethnic Study of Atheroslerosis (MESA) and the Hispanic Community Health Study / Study of Latinos (HCHS/SOL) are available through dbGaP. The dbGaP accession numbers are MESA phs000209 and HCHS/SOL phs000810.

## Acknowledgments

This work was supported by NCCIH R01 AT008621. The Multi-Ethnic Study of Atherosclerosis: MESA and the MESA SHARe project are conducted and supported by the National Heart, Lung, and Blood Institute (NHLBI) in collaboration with MESA investigators. Support for MESA is provided by contracts HHSN268201500003I, N01-HC-95159, N01-HC-95160, N01-HC-95161, N01-HC-95162, N01-HC-95163, N01-HC-95164, N01-HC-95165, N01-HC-95166, N01-HC-95167, N01-HC-95168, N01-HC-95169, UL1-TR-000040, UL1-TR-001079, UL1-TR-001420, UL1-TR-001881, and DK063491. Funding for SHARe genotyping was provided by NHLBI Contract N02-HL-64278. Genotyping was performed at Affymetrix (Santa Clara, California, USA) and the Broad Institute of Harvard and MIT (Boston, Massachusetts, USA) using the Affymetrix Genome-Wide Human SNP Array 6.0. <u>*The Hispanic Community Health Study/Study of Latinos:*</u> HCHS/SOL is a collaborative study supported by contracts from the National Heart, Lung, and Blood Institute (NHLBI) to the University of North Carolina (HHSN268201300001I / N01-HC-65233), University of Miami (HHSN268201300004I / N01-HC-65234), Albert Einstein College of Medicine (HHSN268201300002I / N01-HC-65235), University of Illinois at Chicago (HHSN268201300003I / N01-HC-65236 Northwestern Univ), and San Diego State University (HHSN268201300005I / N01-HC-65237). The following Institutes/Centers/Offices have contributed to the HCHS/SOL through a transfer of funds to the NHLBI: National Institute on Minority Health and Health Disparities, National Institute on Deafness and Other Communication Disorders, National Institute of Dental and Craniofacial Research, National Institute of Diabetes and Digestive and Kidney Diseases (NIDDK), National Institute of Neurological Disorders and Stroke, and NIH Institution-Office of Dietary Supplements. The authors thank the staff and participants of HCHS/SOL for their important contributions. A complete list of staff and investigators has been provided by Sorlie P., et al. in Ann Epidemiol. 2010 Aug;20: 642-649 and is also available on the study website http://www.cscc.unc.edu/hchs/. Other funding sources for this study include R01HL060712, R01HL140976, and R01HL136266 from the NHLBI; and R01DK119268, R01DK120870 and the New York Regional Center for Diabetes Translation Research (P30 DK111022) from the NIDDK. The Genetic Analysis Center at the University of Washington was supported by NHLBI and NIDCR contracts (HHSN268201300005C AM03 and MOD03). Genotyping efforts were supported by NHLBI HSN 26220/20054C, NCATS CTSI grant UL1TR000123, and NIDDK Diabetes Research Center (DRC) grant DK063491. <u>*The Arizona Insulin Resistance Registry:*</u> The AIR registry was supported by Health Research Alliance Arizona and the Center for Metabolic Biology at Arizona State University. Data management support was provided by a grant (UL1 RR024150) from the Mayo Clinic to utilize Research Electronic Data Capture (REDCap).

## Author contributions

CY, BH, JC and AM analyzed the data. CY, BH, JC, TDO, LMR, ACW, MYT, RNL, DKC, LMJ, QQ, IR, SSR, RAM, FHC and AM contributed to interpretation of the data. FHC, AM, RAM and SSR conceptualized and designed the study. CY, BH, AM and FHC wrote the manuscript. All authors contributed to critical editing of the manuscript.

