## Supplemental Text and Figures for "Amerind ancestry predicts the impact of *FADS* genetic variation on omega-3 PUFA deficiency, cardiometabolic and inflammatory risk in Hispanic populations"

### Methods

#### Fatty Acid measurements

The fatty acids were measured in EDTA plasma, frozen at –70°C, using methods previously described by Cao.[1] Lipids were extracted from the plasma using a chloroform/methanol extraction method and the cholesterol esters, triglyceride, phospholipids and free fatty acids are separated by thin layer chromatography. The fatty acid methyl esters were obtained from the phospholipids and were detected by gas chromatography flame ionization. Individual fatty acids were expressed as a percent of total fatty acids. A total of 28 fatty acids were identified. Here, we focus on the following n-3 and n-6 fatty acids: eicosapentaenoic acid (EPA), docosapentaenoic acid (DPA), docosahexaenoic acid (DHA), and arachidonic acid (ARA).

#### Dietary fish intake and supplement use in MESA

At the baseline examination, usual dietary intake over the previous year was assessed with a modified Block-style 120-item Food Frequency Questionnaire (FFQ).[2,3] Fish intake was quantified as the sum of the servings per day for the following food categories: fried fish, shrimp, tuna, boiled fish, fish stew and stir-fried shrimp. Use of nutritional supplements including "Cod liver oil, other fish oils or omega-3 fatty acids" was also reported in the MESA FFQ; we used these responses to verify that none of the participants included in our analyses reported use of these supplements.

#### Additional phenotypes in MESA

Fasting blood samples were drawn, processed and stored using standardized procedures. Total cholesterol, HDL- cholesterol and triglycerides were measured at the Collaborative Studies Clinical Laboratory at Fairview-University Medical Center (Minneapolis, MN). Total cholesterol was measured using a cholesterol oxidase method (Roche Diagnostics; Indianapolis, IN), HDL-cholesterol using the cholesterol oxidase method (Roche Diagnostics) after precipitation of non-

HDL-cholesterol with magnesium/dextran, triglycerides using Triglyceride GB reagent (Roche Diagnostics). All lipid measurements were performed on the Roche COBAS FARA centrifugal analyzer.[4]

Height was measured to the nearest 0.1 cm with the subject in stocking feet and weight was measured to the nearest pound with the subject in light clothing using a balanced scale. Waist circumference (WC), and hip circumference were measured to the nearest 0.5 kg, 0.1 cm, and 0.1 cm, respectively.

Plasma soluble ICAM-1 (sICAM-1), and soluble E-selectin (sE-selectin) concentrations were measured in blood samples collected at baseline and were processed with the use of a standardized protocol based on that used in the Cardiovascular Health Study[5] and stored at –80 °C until analyzed at the University of Vermont (Burlington, VT).[6]

**Detection of outliers in measured PUFA levels and selected proximal traits**

Outliers for all of the quantitative traits examined by genetic association analysis were identified by the limits of median +/- 3.5 * MAD', where MAD' is computed with a scale factor constant of 1.4826 [default for the mad() function in R]. The value of MAD' = 1.4826 * $MAD_0$ where $MAD_0$ is the raw value of median absolute deviation. For the all of PUFA traits, as well as the selected lipid levels, anthopometric traits, and inflammatory markers, outliers were winsorized to the value of (median +/- 3.5 * MAD').

**Genotyping and imputation in MESA**

Participants in the MESA cohort who consented to genetic analyses and data sharing (dbGaP) were genotyped using the Affymetrix Human SNP Array 6.0 (GWAS array) as part of the NHLBI CARe (Candidate gene Association Resource) and SHARe (SNP Health Association Resource) projects. Genotype quality control for these data included filter on SNP level call rate < 95%, individual level call rate < 95%, heterozygosity > 53%, described previously[7]. The cleaned genotypic data was deposited with MESA phenotypic data into dbGaP (study accession phs000209.v13.p3); 8,224 consenting individuals (2,685 White, 2,588 non-Hispanic African-American, 2,174 Hispanic, 777 Chinese) were included, with 897,981 SNPs passing study specific quality control (QC). SNP coverage from the original GWAS SNP genotyping array was increased through imputation using the 1,000 Genomes Phase 3 integrated variant set completed using the Michigan Imputation Server (https://imputationserver.sph.umich.edu).

**Genetic association analysis**

Within the n-3 and n-6 LC-PUFA study, outliers were identified using a cutoff based on the median absolute deviation (MAD) and winsorized the selected extreme values for each PUFA level **(Figure S5 and S6).** In analysis of PUFA levels, we included covariate adjustment for age, sex, study site, principal components of ancestry, batch for fatty acid measurement and *FADS* cluster SNP rs174537. ProbABELv0.5.0 [8] was used to conduct genetic association analysis using linear models with robust standard errors. In addition, genetic association results were filtered based on minor allele frequency > 0.05 and imputation R squared > 0.5.

Local association analysis was performed for the 500 kb regions from starting position 61.3 Mb to ending position 61.8 Mb on chromosome 11, and local association plots were generated using LocusZoom [9]. Statistical significance for local association analysis was determined using a Bonferroni-corrected *P*-value with a family-wise error rate $\alpha$=0.05, correcting for the total number 1081 of genetic variants passing filters in the region of interest. Thus, our genetic association analyses were subject to a Bonferroni corrected threshold of $P^*=0.05 / 1081=4.6\times10^{-5}$.

**Global Ancestry Analysis**

Individuals from all of the the1000 Genomes phase 3[10] and the Human Genome Diversity Project (HGDP)[11,12] groups listed below were included in global ancestry analysis in order to inform labels for the inferred ancestry groups.

*1000 Genomes populations:*

- Amerind: MXL (Mexican Ancestry from Los Angeles, USA), PUR (Puerto Ricans from Puerto Rico), CLM (Colombians from Medellin, Colombia) and PEL (Peruvians from Lima, Peru) were used to represent Amerind populations from the 1000 Genomes.
- African: ASW (Americans of African Ancestry in SW, USA), ACB (African Caribbeans in Barbados), ESN (Esan in Nigeria); GWD (Gambian in Western Divisions in the Gambia); LWK (Luhya in Webuye, Kenya), MSL (Mende in Sierra Leone) and YRI (Yoruba in Ibadan, Nigeria) groups were used to represent African populations from the 1000 Genomes.
- European: Utah Residents (CEPH) with Northern and Western European Ancestry (CEU), Toscani in Italia (TSI), Finnish in Finland (FIN), British in England and Scotland (GBR), Iberian Population in Spain (IBS) set were used to represent the non-admixed European population from the 1000 Genomes.

*Human Genome Diversity Project (HGDP):*

- Amerind: Colombian, Karaitiana, Maya, Pima and Surui were used to represent Amerind populations in the HGDP.
- African: Bantu, Biaka, Mandenka, Mbuti pygmy, Mozabite, San and Yoruba were used to represent African populations in the HGDP.
- European: Adygei, Basque, French, North Italian, Orcadian, Russian, Sardinian and Tuscan were used to represent the non-admixed European population in the HGDP.

Model-based cluster analysis was applied to estimate proportions of European, African and Amerind ancestry for all genotyped individuals using the package ADMIXTURE[14]. This software package used a block relaxation approach implemented with a novel quasi-Newton acceleration method that makes the method computationally feasible for much larger data sets, both in terms of the number of individuals and the number of genetic variants.

**Local Ancestry Analysis**

In order to construct an appropriate reference panel for local ancestry analysis, we sought to identify representative non-admixed Amerind, African and European individuals from among the 1000 Genomes phase 3[10] and the HGDP[11,12] groups included in global ancestry analysis, as noted above. As previous research has noted a high degree of admixture among some of the 1000 Genomes Amerind populations[13], we initially examined the mean global proportion of Amerind ancestry in each of the putative Amerind groups from the 1000 Genomes and the HGDP. Similar to the previous report[13], we noted the overall global proportion of Amerind ancestry was <0.5 in each of MXL, PUR and CLM from the 1000 Genomes. Thus, we removed these groups from further consideration. We confirmed that the remaining 1000 Genomes PEL group, as well as all of the HGDP Amerind groups exhibited mean global Amerind ancestry > 0.7. Thus, these groups were carried forward for further consideration. We further cleaned the remaining 1000 Genomes and HGDP reference samples for each of the continental super-populations, by applying a threshold for each population, such that inclusion in the Amerind, African or European reference panel used for local ancestry analysis required at least 90%, 99% or 99% global proportions of ancestry for the respective continent represented. After applying this filter, there were 36, 371 and 434 individuals included in the resulting reference panels for Amerind, African and European ancestry, respectively.

Local ancestry was defined as the genetic ancestry of each individual for the region of *FADS* SNP rs174537, where each individual can have 0, 1 or 2 copies of an allele derived from each ancestral population. Local ancestry, was estimated using the RFMix package.[15] This tool implements a discriminative modeling approach by dividing the region on chromosome 11 into windows. Local ancestry is inferred within each window by using a conditional random field (CRF), parameterized by random forest trained using reference data representing the selected continental source populations (European, Amerind and African).

**Follow-up in the AIR and HCHS/SOL cohorts**

*Arizona Insulin Resistance (AIR) registry:* The AIR registry comprises 667 individuals (aged 8-83 years) and has been described in detail elsewhere.[16] Within the AIR registry, there are 497 adult Hispanic participants (18-83 years) who agreed and consented to have their samples stored and analyzed for future projects (such as this study). Genotyping of *FADS* rs174537 was performed using a TaqMan SNP genotyping assay. Anthropometric data for the AIR registry participants include weight, height, body mass index, % body fat, hip circumference, and waist circumference. The cardio-metabolic measures obtained from each participant included glucose, hemoglobin A1C, insulin, triglycerides, total cholesterol, high-density lipoprotein cholesterol (HDL), low-density lipoprotein cholesterol (LDL) and very-low-density lipoprotein cholesterol (VLDL).

*Hispanic Community Health Study (HCHS/SOL):* The HCHS/SOL is a prospective population-based study of 16,415 Hispanic/Latino adults aged 18 to 74 years at recruitment who were living in four US metropolitan areas (Bronx, NY; Chicago, IL; Miami, FL; and San Diego, CA).[17,18] A comprehensive battery of interviews and a clinical assessment with blood draw (both fasting and two hours after a 75 gram glucose load) were conducted at in-person clinic visits during 2008 to 2011 (Visit 1 baseline) and during 2014 to 2017 (Visit 2). Information on demographics, behaviors, health status, family and medical histories, and medication use was collected using structured questionnaires, and blood pressure and anthropometric traits were measured. Blood biomarkers including blood lipids and glycemic traits, were measured by standard methods.[19] Annual follow-up telephone interviews were conducted to ascertain information on health status. The study was approved by the institutional review boards at all participating institutions, and all participants gave written informed consent.

Genotyping was performed with an Illumina custom array (15041502 B3), which consists of the Illumina Omni 2.5M array (HumanOmni2.5-8v1-1) plus ~150k custom SNPs, with the quality control (QC) performed at HCHS/SOL Genetic Analysis Center. The variant rs174537 was genotyped directly through this effort in HCHS/SOL. An iterative procedure was used to simultaneously estimate principal components (PCs) reflecting population structure and kinship coefficients measuring familial relatedness.[20–22]

**Table S1: Regression of n-3 and n-6 PUFAs on global ancestry, also accounting for local ancestry and rs174537 SNP genotype.**

| | Columbia University | | UCLA | | University of Minnesota | | Meta Analysis | |
|---|---|---|---|---|---|---|---|---|
| **Model 1** | | | | | | | | |
| | BETA | P-value | BETA | P-value | BETA | P-value | BETA | P-value |
| EPA (%) | -0.22 | 0.097 | -0.38 | $1.6 \times 10^{-05}$ | -0.65 | $1.7 \times 10^{-14}$ | -0.48 | $3.7 \times 10^{-23}$ |
| DPA (%) | -0.14 | 0.086 | -0.14 | 0.036 | -0.27 | $1.4 \times 10^{-04}$ | -0.18 | $7.6 \times 10^{-06}$ |
| DHA (%) | -0.34 | 0.402 | -0.33 | 0.325 | -0.91 | $6.3 \times 10^{-04}$ | -0.63 | 0.0007 |
| ARA (%) | -4.86 | $9.8 \times 10^{-07}$ | -4.08 | $3.2 \times 10^{-07}$ | -3.45 | $4.6 \times 10^{-05}$ | -4.06 | $1.3 \times 10^{-16}$ |
| **Model 2** | | | | | | | | |
| | Columbia University | | UCLA | | University of Minnesota | | Meta Analysis | |
| | BETA | P-value | BETA | P-value | BETA | P-value | BETA | P-value |
| EPA (%) | -0.05 | 0.702 | -0.16 | 0.067 | -0.51 | $5.5 \times 10^{-09}$ | -0.28 | $3.7 \times 10^{-08}$ |
| DPA (%) | -0.01 | 0.873 | 0.02 | 0.792 | -0.18 | 0.017 | -0.05 | 0.185 |
| DHA (%) | 0.09 | 0.844 | -0.05 | 0.883 | -0.71 | 0.011 | -0.37 | 0.056 |
| ARA (%) | -1.47 | 0.084 | -0.75 | 0.287 | -0.64 | 0.372 | -0.89 | 0.038 |
| **Model 3** | | | | | | | | |
| | Columbia University | | UCLA | | University of Minnesota | | Meta Analysis | |
| | BETA | P-value | BETA | P-value | BETA | P-value | BETA | P-value |
| EPA (%) | 0.08 | 0.570 | -0.12 | 0.179 | -0.48 | $3.1 \times 10^{-08}$ | -0.34 | $8.4 \times 10^{-07}$ |
| DPA (%) | 0.04 | 0.683 | 0.03 | 0.653 | -0.14 | 0.059 | -0.07 | 0.192 |
| DHA (%) | 0.20 | 0.696 | 0.06 | 0.879 | -0.69 | 0.018 | -0.47 | 0.058 |
| ARA (%) | -1.16 | 0.199 | -0.39 | 0.583 | -0.08 | 0.914 | -0.52 | 0.364 |

**Table S1** shows the regression analysis results for Model 1-3 stratified by study site and combined by meta-analysis. All beta estimates for effects of the global ancestry on fatty acid levels are presented in units of percent total fatty acids.

Model 1. PUFA ~ age + sex + fish intake + global proportions of Amerind ancestry

Model 2. PUFA ~ age + sex + fish intake + global proportions of Amerind ancestry + rs174537 genotype

Model 3. PUFA ~ age + sex + fish intake + global proportions of Amerind ancestry + rs174537 genotype + local proportion of Amerind ancestry

**Table S2: Interaction analysis among rs174537, local and global ancestry**

| | Model 1 | | | | | | | |
|---|---|---|---|---|---|---|---|---|
| | Columbia University | | UCLA | | University of Minnesota | | Meta Analysis | |
| | BETA | P-value | BETA | P-value | BETA | P-value | BETA | P-value |
| EPA (%) | -0.29 | 0.154 | 0.05 | 0.643 | 0.15 | 0.184 | 0.05 | 0.496 |
| DPA (%) | -0.04 | 0.778 | -0.04 | 0.672 | -0.04 | 0.662 | -0.04 | 0.503 |
| DHA (%) | 0.33 | 0.649 | -0.03 | 0.958 | 0.02 | 0.948 | 0.05 | 0.852 |
| ARA (%) | -0.09 | 0.939 | 0.46 | 0.624 | -1.71 | 0.075 | -0.49 | 0.4001 |
| | Model 2 | | | | | | | |
| | Columbia University | | UCLA | | University of Minnesota | | Meta Analysis | |
| | BETA | P-value | BETA | P-value | BETA | P-value | BETA | P-value |
| EPA (%) | -0.06 | 0.265 | -0.02 | 0.328 | 0.01 | 0.769 | -0.02 | 0.366 |
| DPA (%) | -0.003 | 0.927 | -0.03 | 0.165 | -0.05 | 0.118 | -0.03 | 0.06 |
| DHA (%) | 0.17 | 0.417 | 0.13 | 0.289 | 0.06 | 0.587 | 0.105 | 0.177 |
| ARA (%) | 0.13 | 0.720 | -0.18 | 0.457 | -0.39 | 0.194 | -0.178 | 0.279 |

**Table S2** shows the regression analysis results for interaction effect among rs174537, local and global ancestry (Model 1-2) stratified by study site and combined by meta-analysis. All beta estimates for interaction effects on fatty acid levels are presented in units of percent total fatty acids.
Model 1. PUFA ~ age + sex + fish intake + global proportions of Amerind ancestry + rs174537 genotype + local proportion of Amerind ancestry + rs174537 genotype * global proportion of Amerind ancestry
Model 2. PUFA ~ age + sex + fish intake + global proportions of Amerind ancestry + rs174537 genotype + local proportion of Amerind ancestry + rs174537 genotype * local proportion of Amerind ancestry

**Table S3: Genotypic effects of rs174537 on Fasting Lipids, Anthropometrics and Inflammatory Traits.**

| | | | Model 1 | | Model 2 | |
|---|---|---|---|---|---|---|
| | | | Beta | P value | Beta | P value |
| Fasting Lipids | Triglycerides (mg/dL) | GT | 21.27 | 0.0001 | 11.81 | 0.044 |
| | | TT | 29.94 | $1.01 \times 10^{-06}$ | 16.19 | 0.018 |
| | HDL-C (mg/dL) | GT | -1.30 | 0.141 | -0.35 | 0.708 |
| | | TT | -2.48 | 0.010 | -0.95 | 0.384 |
| Anthropometrics | waist-hip ratio | GT | 0.006 | 0.152 | -0.002 | 0.710 |
| | | TT | 0.013 | $8.94 \times 10^{-03}$ | -0.0007 | 0.908 |
| | Height (cm) | GT | -1.36 | 0.002 | -0.39 | 0.412 |
| | | TT | -3.46 | $6.59 \times 10^{-12}$ | -1.62 | 0.003 |
| | Weight (kg) | GT | -1.89 | 0.077 | -1.36 | 0.230 |
| | | TT | -3.12 | $7.48 \times 10^{-03}$ | -2.41 | 0.067 |
| | BMI (kg/m^2) | GT | -0.25 | 0.50 | -0.39 | 0.33 |
| | | TT | -0.002 | 0.99 | -0.37 | 0.42 |
| Inflammatory | s-ICAM (ng/mL) | GT | 30.64 | 0.002 | 19.09 | 0.081 |
| | | TT | 26.09 | 0.018 | 10.22 | 0.411 |
| | e-Selectin (ng/mL) | GT | 10.00 | 0.048 | 10.41 | 0.051 |
| | | TT | 11.50 | 0.032 | 10.49 | 0.080 |

**Table S3** shows the regression analysis results for the effect of rs174537 genotype compaed to two copies of the reference allele G on inflammatory, fasting Lipids and anthropometrics with adjustment for age and sex (Model 1) and Model 1 + first four principal components of ancestry (Model 2).

**Table S4: Genotypic effects of rs174557 on Fasting Lipids, Anthropometrics and Inflammatory Traits.**

| | | | Model 1 | | Model 2 | |
|---|---|---|---|---|---|---|
| | | | Beta | P value | Beta | P value |
| Fasting Lipids | Triglycerides (mg/dL) | AG | 22.92 | $2.55 \times 10^{-05}$ | 14.21 | 0.012 |
| | | GG | 29.94 | $9.56 \times 10^{-07}$ | 16.99 | 0.012 |
| | HDL-C (mg/dL) | AG | -1.48 | 0.085 | -0.59 | 0.513 |
| | | GG | -2.40 | 0.012 | -0.93 | 0.390 |
| Anthropometrics | waist-hip ratio | AG | 0.008 | 0.095 | -0.0004 | 0.932 |
| | | GG | 0.013 | 0.008 | -0.0002 | 0.972 |
| | Height (cm) | AG | -1.66 | 0.0001 | -0.747 | 0.103 |
| | | GG | -3.84 | $1.57 \times 10^{-14}$ | -2.079 | $1.48 \times 10^{-04}$ |
| | Weight (kg) | AG | -4.18 | 0.068 | -2.95 | 0.219 |
| | | GG | -7.46 | 0.003 | -6.04 | 0.036 |
| | BMI (kg/m^2) | AG | -0.14 | 0.64 | -0.25 | 0.522 |
| | | GG | 0.05 | 0.91 | -0.32 | 0.483 |
| Inflammatory | s-ICAM (ng/mL) | AG | 30.24 | 0.002 | 20.02 | 0.058 |
| | | GG | 26.14 | 0.017 | 11.54 | 0.343 |
| | e-Selectin (ng/mL) | AG | 7.29 | 0.139 | 7.21 | 0.161 |
| | | GG | 8.83 | 0.101 | 6.93 | 0.239 |

**Table S4** shows the regression analysis results for the effect of rs174557 genotype compared to two copies of the reference allele A on inflammatory, fasting Lipids and anthropometrics with adjustment for age and sex (Model 1) and Model 1 + first four principal components of ancestry (Model 2).

**Table S5. Participant Characteristics in AIR registry.**

| Characteristics | N = 497 |
|---|---|
| Sex (Female) | 64.60% |
| Age (Years) | 36.4 (11.2) |
| Height (cm) | 163 (8.70) |
| Weight (kg) | 80.8 (19.5) |
| HDL-C (mg/dL) | 43.9 (11.2) |
| LDL-C (mg/dL) | 106.6(28.6) |
| Triglycerides (mg/dL) | 136.6 (79.7) |
| Total Cholesterol (mg/dL) | 173.9 (34.9) |
| Body mass index (kg/m^2) | 29.9 (5.8) |
| Waist-to-Hip ratio | 0.9 (0.1) |

Table S5 shows phenotypic descriptive statistics that are presented as percentages for dichotomous variables and mean (standard deviation) for continuous variables.

**Table S6. Participant Characteristics in HCHS/SOL cohort.**

| Characteristics | Self-reported Hispanic country/region of origin | | | | | | Total (N=12333) |
|---|---|---|---|---|---|---|---|
| | **Cuba** (N=2190) | **Dominican** (N=1157) | **PuertoRico** (N=2178) | **SouthAmer** (N=899) | **CentralAmer** (N=1335) | **Mexico** (N=4537) | |
| Sex (Female) | (53.8%) | (65.7%) | (58.3%) | (60.2%) | (60.1%) | (60.9%) | (59.5%) |
| Age (years) | 48.7 (13.3) | 45.5 (14.3) | 47.7 (14.3) | 46.4 (13.3) | 44.5 (13.4) | 44.6 (13.9) | 46.1 (13.9) |
| Height (cm) | 164.1 (9.2) | 162 (8.9) | 163 (9.4) | 160.8 (9.3) | 160 (8.7) | 161.3 (9.4) | 162.0 (9.3) |
| Weight (kg) | 78.9 (17.7) | 77.4 (16.5) | 82 (20.1) | 74 (15.5) | 76.9 (16.6) | 77.8 (17.8) | 78.4 (17.9) |
| BMI (kg/m^2) | 29.3 (5.9) | 29.5 (5.8) | 30.9 (7) | 28.6 (5.2) | 30 (5.8) | 29.9 (6) | 29.8 (6.1) |
| Waist-hip ratio | 0.9 (0.1) | 0.9 (0.1) | 0.9 (0.1) | 0.9 (0.1) | 0.9 (0.1) | 0.9 (0.1) | 0.9 (0.1) |
| HDL (mg/dl) | 49 (12.9) | 51.6 (13) | 49.5 (13.5) | 50.4 (13.2) | 48.6 (12.9) | 49.2 (12.6) | 49.5 (13.0) |
| LDL (mg/dl) | 130.1 (38) | 121.3 (38.8) | 115.9 (35.6) | 126.1 (35.5) | 124 (35.8) | 122.2 (35.4) | 122.9 (36.6) |
| Triglycerides (mg/dl) | 134.6 (71.2) | 107.3 (57.7) | 122.6 (66.4) | 131.7 (71.5) | 142.6 (72.7) | 132.9 (66.9) | 129.8 (68.4) |
| Total cholesterol (mg/dl) | 206 (44) | 194.4 (43.2) | 189.8 (41.5) | 203 (41.6) | 201.1 (41.7) | 198 (41.1) | 198.3 (42.3) |
| C-Reactive orotein (CRP) (mg/L) | 4.3 (6.6) | 3.9 (6.6) | 4.7 (6.9) | 3.4 (7.7) | 4.3 (10.3) | 3.8 (7.3) | 4.1 (7.5) |
| Effect Allele Frequency of rs174537 (effect allele: T) | 0.28 | 0.26 | 0.38 | 0.63 | 0.61 | 0.63 | 0.48 |

Table S6 shows phenotypic descriptive statistics that are presented as percentages for dichotomous variables and mean (standard deviation) for continuous variables.

**Table S7. Analysis of rs174537 effects on triglycerides and waist-hip ratio in AIR registry and HCHS/SOL.**

| | AIR cohort | | HCHS/SOL cohort | |
|---|---|---|---|---|
| | Beta | P-value | Beta | P-value |
| Triglycerides | 10.39 | 0.03 | 8.75 | $5.84 \times 10^{-25}$ |
| Waist-hip ratio | 0.01 | 0.20 | 0.003 | $2.77 \times 10^{-05}$ |
| Height | -0.49 | 0.20 | -1.33 | $4.47 \times 10^{-56}$ |
| Weight | 0.43 | 0.71 | -1.25 | $2.61 \times 10^{-08}$ |

**Table S7** shows the additive effect of *FADS* rs174537 allele T on Triglycerides, waist-hip ratio, height and weight in the AIR registry and the HCHS/SOL cohort. The effect estimates are adjusted for age and sex in both studies (plus additional random-effects adjustment for household block and unit sharing in HCHS/SOL).

**Table S8: Stratified analysis of rs174537 effects on triglycerides and waist-hip ratio HCHS/SOL, with additional adjustment for principal components of ancestry.**

| | Chicago | | Miami | | San Diego | | Bronx | | Meta Analysis | |
|---|---|---|---|---|---|---|---|---|---|---|
| | BETA | P-value | BETA | P-value | BETA | P-value | BETA | P-value | BETA | P-value |
| Triglycerides | 4.94 | 0.015 | 2.85 | 0.13 | 4.47 | 0.014 | 4.04 | 0.019 | 4.05 | $1.26x10^{-05}$ |
| Waist-hip ratio | -0.0005 | 0.79 | -0.003 | 0.096 | -0.0008 | 0.63 | -0.003 | 0.19 | -0.002 | 0.068 |
| Height | 0.006 | 0.975 | -0.035 | 0.834 | -0.163 | 0.354 | -0.33 | 0.06 | -0.129 | 0.138 |
| Weight | -0.034 | 0.943 | 0.293 | 0.518 | 0.465 | 0.369 | -1.02 | 0.038 | -0.068 | 0.778 |

**Table S8** shows the genotypic effect of *FADS* SNP rs174537 on triglycerides, waist-hip ratio, height and weight in the HCHS/SOL cohort followed by a fixed-effects meta-analysis. The effect estimates are adjusted for age, sex, study site, Hispanic region of origin, first 5 principal components of ancestry, household block and unit sharing.

**Table S9: Regression of n-3 and n-6 PUFAs on Study Site in MESA Hispanic Participants**

| | | BETA | P-value |
|---|---|---|---|
| EPA (% of total LC-PUFA) | UCLA | -0.22 | $3.81 \times 10^{-20}$ |
| | University of Minnesota | -0.19 | $8.72 \times 10^{-21}$ |
| DPA (% of total LC-PUFA) | UCLA | -0.06 | 0.0003 |
| | University of Minnesota | -0.08 | $3.10 \times 10^{-09}$ |
| DHA (% of total LC-PUFA) | UCLA | -1.34 | $6.36 \times 10^{-54}$ |
| | University of Minnesota | -1.02 | $2.25 \times 10^{-41}$ |
| ARA (% of total LC-PUFA) | UCLA | -1.55 | $2.39 \times 10^{-15}$ |
| | University of Minnesota | -1.37 | $2.60 \times 10^{-15}$ |

**Table S9** shows the study-site effects on LC-PUFA levels are reported based on assignment of Columbia University as the reference group for estimation of site-specific effects. Results are based on covariate adjustment for age and sex.

**Figure S1: Relationship of LC-PUFA levels with Global Proportion of Amerind Ancestry before and after adjustment for rs174557 genotype.**

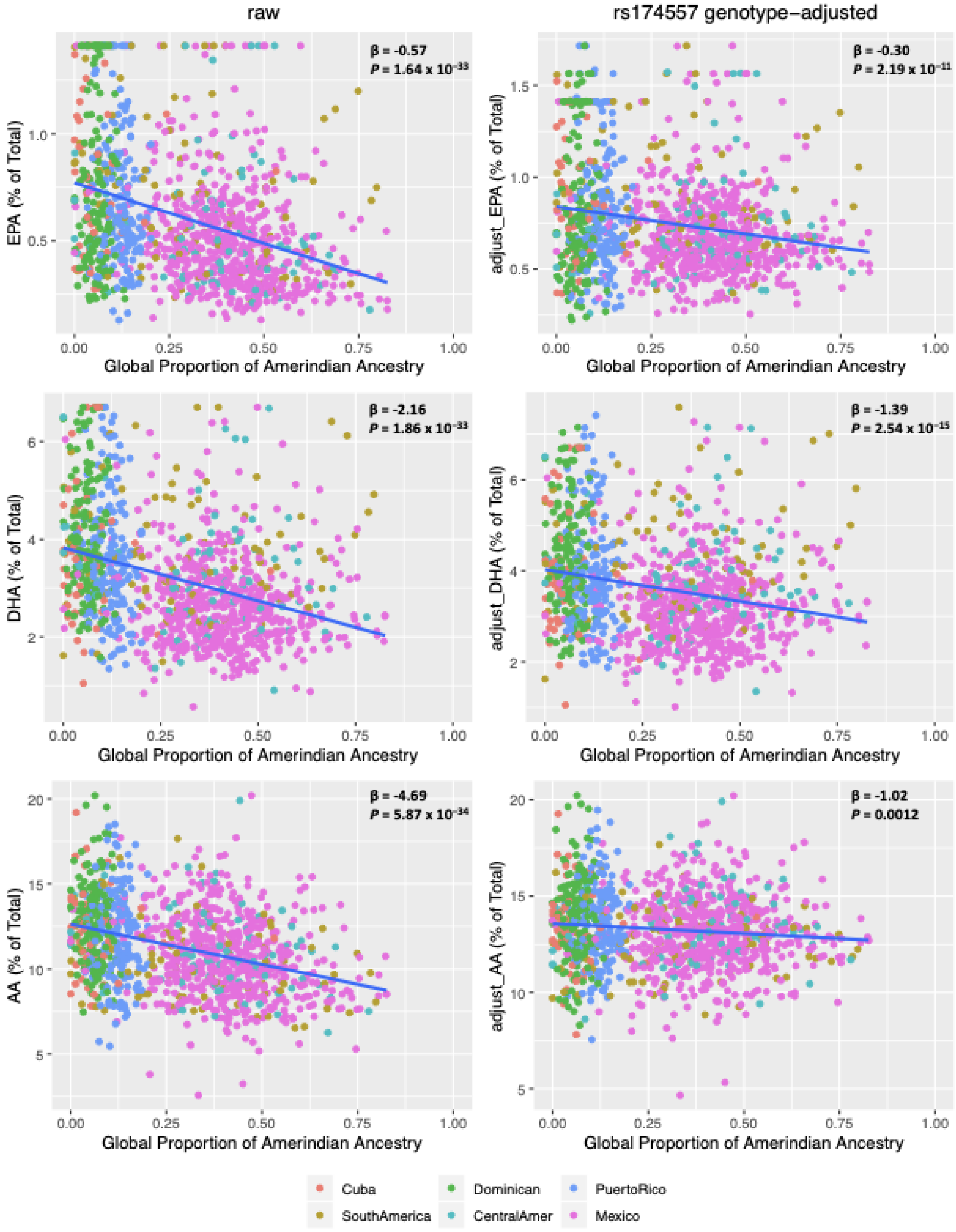


The regression effect estimates ($\beta$ expressed as % of total fatty acids) and *P*-values are shown in the upper right corner of each panel. **Left** panels show the relationship of raw LC-PUFA levels with Global Proportion of Amerind Ancestry as estimated from genome-wide SNP data. **Right** panels show the relationship of rs174557 genotype-adjusted LC-PUFA levels with Global Proportion of Amerind Ancestry. Here, the adjusted LC-PUFA levels were obtained as residuals after regression against rs174557 genotype and re-centered around the raw mean.

**Figure S2**: **Relationship of triglycerides with Global Proportion of Amerind Ancestry before and after adjustment for rs174557 genotype.**

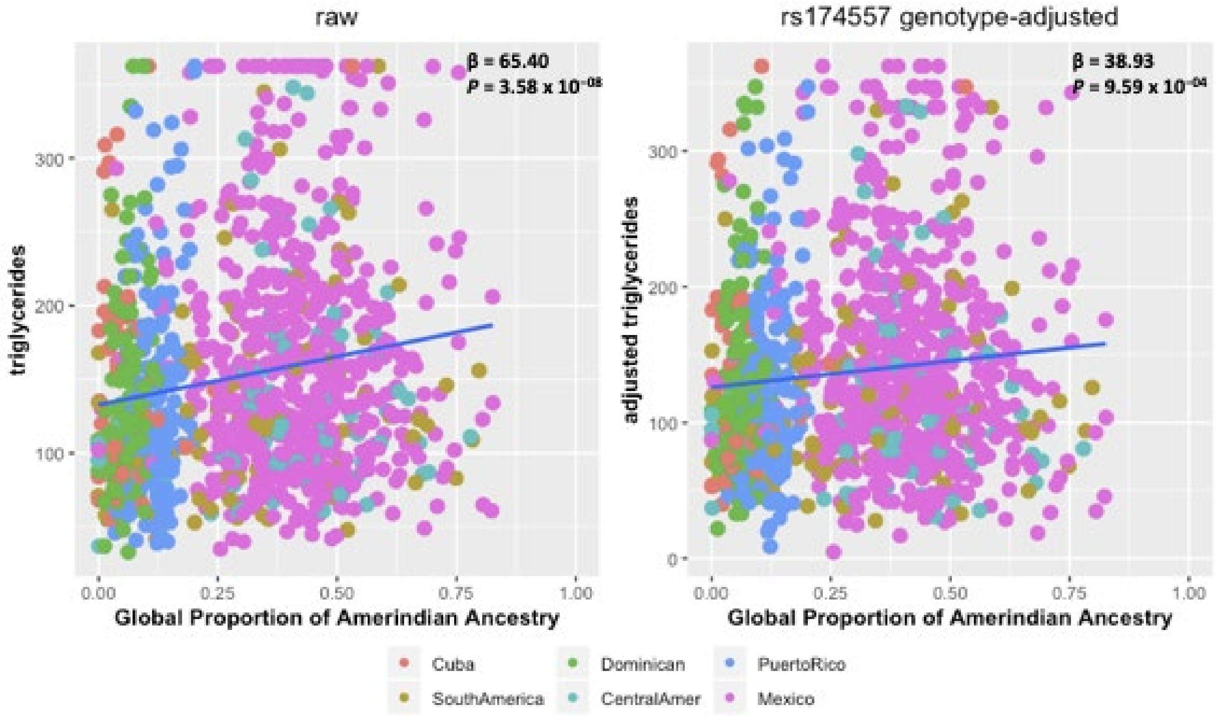


The regression effect estimates ($\beta$ in mg/dL) and *P*-values are shown in the upper right corner of each panel. **Left** figure shows the relationship of raw triglyceride levels with Global Proportion of Amerind Ancestry. **Right** figure shows the relationship of rs174557 genotype-adjusted triglyceride levels with Global Proportion of Amerind Ancestry. Here, rs174557 genotype-adjusted triglyceride levels were obtained as residuals from regression accounting for rs174557 genotype, and re-centered around the raw means.

**Figure S3: Conditional local association plots for n-3 and n-6 PUFAs in the *FADS1/2* region, accounting for the rs174537 SNP**

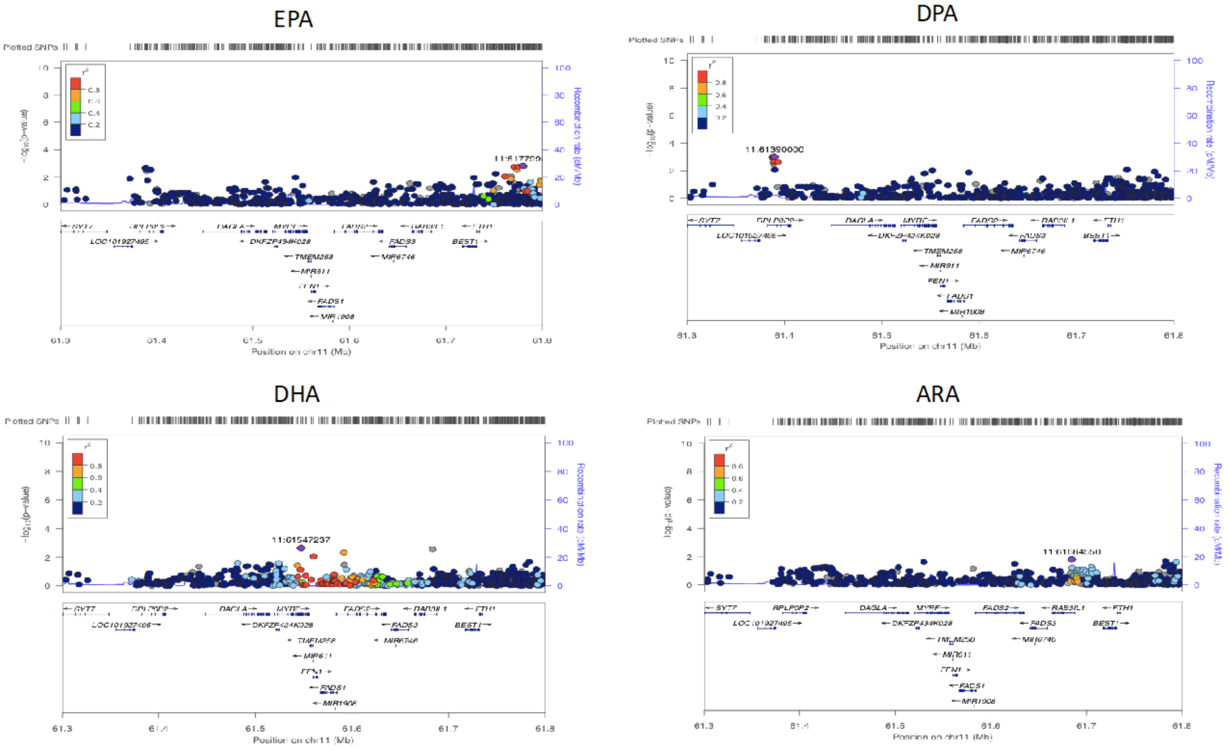


**Figure S3** shows the conditional local association results for n-3 and n-6 PUFAs in the FADS1/2 region, accounting for the rs174537 SNP.

**Figure S4: Genotypic effects of rs174537 on triglycerides and E-selectin.**

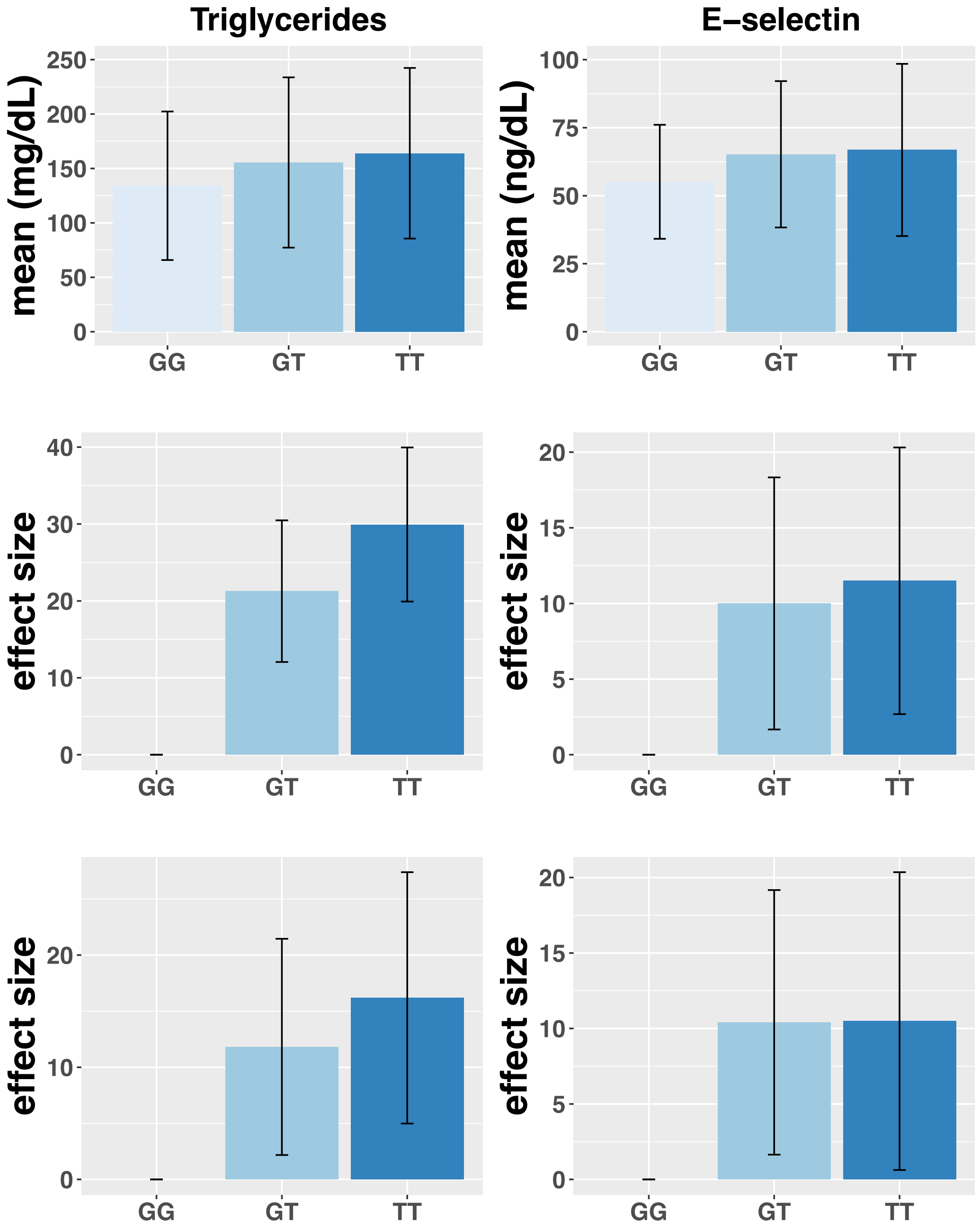


**Figure S4** shows the effect of rs174537 on triglycerides and E-selectin. The sample size across genotype is 293 for GG, 484 for GT, 325 for TT. Upper figure shows the mean and standard deviation of Triglycerides and E-selectin stratified by the genotypes of rs174537. Center figure shows the estimated effect and standard error of carrying one or two copies of the ancestral allele (compared to the reference of zero). The effect-size estimates are adjusted for age and sex. Bottom figure shows the estimated effect and standard error of carrying one or two copies of the ancestral allele (compared to a reference of zero), adjusted for age, sex and principal components of ancestry.

**Figure S5: Distribution of n-3 and n-6 PUFAs in MESA Hispanic participants.**

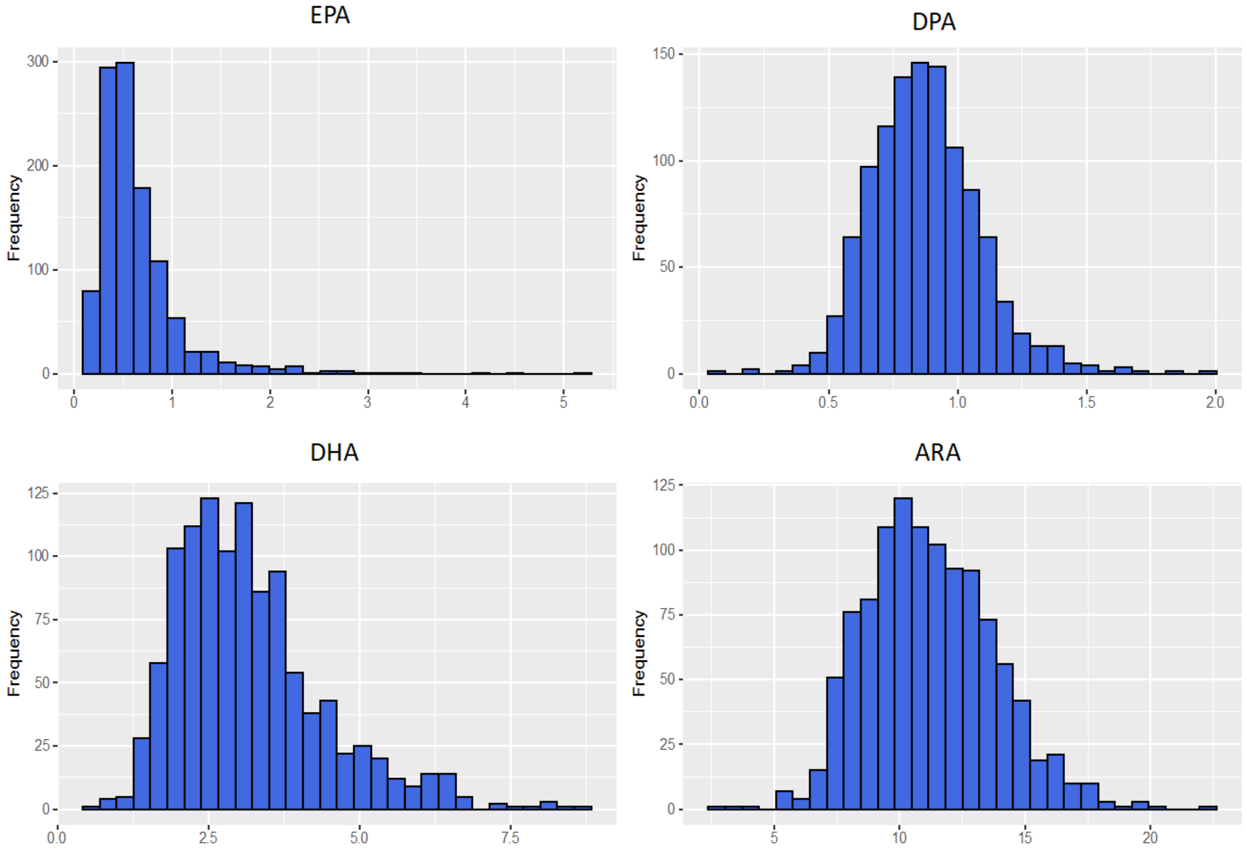


**Figure S5** shows the distribution of n-3 and n-6 PUFAs in MESA Hispanic participants.

**Figure S6 Distribution of n-3 and n-6 PUFAs in MESA Hispanic participants after winsorizing at median +/- 3.5 Median Absolute Deviation (MAD)**

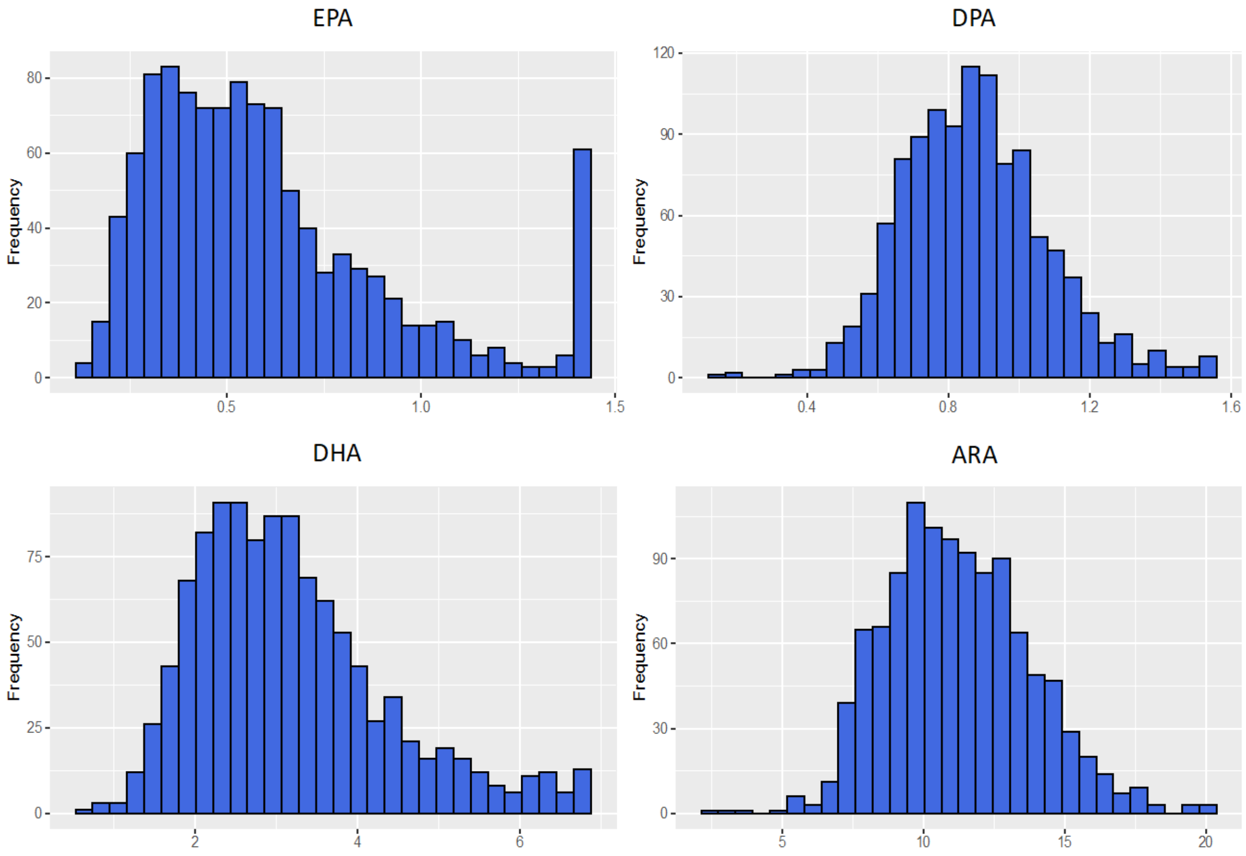


**Figure S6** shows the distribution of n-3 and n-6 PUFAs in MESA Hispanic participants after winsorizing at median +/- 3.5 Median Absolute Deviation (MAD)
